# *Legionella pneumophila* occurrence in reduced-occupancy buildings in 11 cities during the COVID-19 pandemic

**DOI:** 10.1101/2022.06.28.22277022

**Authors:** Katherine S. Dowdell, Hannah D. Greenwald, Sayalee Joshi, Marianne Grimard-Conea, Sarah Pitell, Yang Song, Christian Ley, Lauren C. Kennedy, Solize Vosloo, Linxuan Huo, Sarah-Jane Haig, Kerry A. Hamilton, Kara L. Nelson, Ameet Pinto, Michele Prévost, Caitlin R. Proctor, Lutgarde M. Raskin, Andrew J. Whelton, Emily Garner, Kelsey J. Pieper, William J. Rhoads

**Author notes:** <u>**Corresponding Authors**</u> William J. Rhoads – Department of Environmental Microbiology, Swiss Federal Institute of Aquatic Science and Technology (EAWAG), Dubendorf, Switzerland;., Emily Garner – Wadsworth Department of Civil and Environmental Engineering, West Virginia University, Morgantown, WV, 26506, USA;., Kelsey J. Pieper – Department of Civil and Environmental Engineering, Northeastern University, Boston, MA, 02115, USA;. Co-first authors (Katherine S. Dowdell, Hannah D. Greenwald).

## Abstract

In spring 2020, reduced water demand was an unintended consequence of COVID-19 pandemic-related building closures. Concerns arose that contaminants associated with water stagnation, such as *Legionella pneumophila*, could become prevalent. To investigate this potential public health risk, samples from 26 reduced-occupancy buildings across 11 cities in the United States, Canada, and Switzerland were analyzed for *L. pneumophila* using liquid culture (Legiolert, n=258) and DNA-based methods (qPCR/ddPCR, n=138). *L. pneumophila* culture-positivity was largely associated with just five buildings, each of which had specific design or operational deficiencies commonly associated with *L. pneumophila* occurrence. Samples from free chlorine buildings had higher culture-positivity (37%) than chloramine buildings (1%), and 78% of culture-positive samples occurred when the residual was ≤0.1 mg/L Cl_2_. Although overall sample positivities using culture- and DNA-based methods were equivalent (34% vs. 35%), there was disagreement between the methods in 13% of paired samples. Few buildings reported any water management activities, and *L. pneumophila* concentrations in flushed samples were occasionally greater than in first-draw samples. This study provides insight into how building plumbing characteristics and management practices contribute to *L. pneumophila* occurrence during low water use periods and can inform targeted prevention and mitigation efforts.

**Synopsis Statement:** *Legionella pneumophila* occurrence was evaluated in reduced-occupancy buildings during the COVID-19 pandemic across multiple cities.

**Graphic for Table of Contents (TOC):** 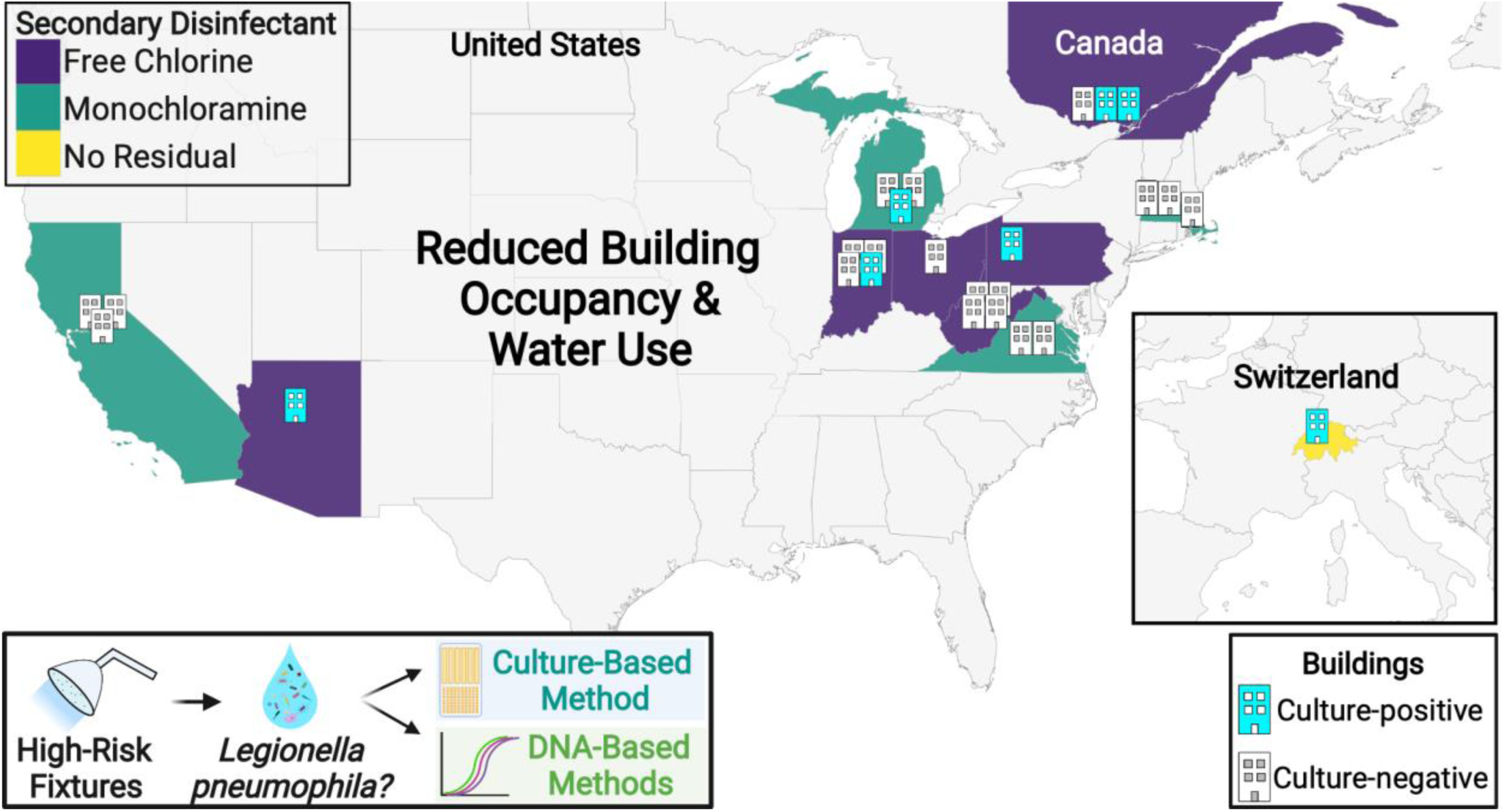

## 1. Introduction

Closures of non-residential buildings to slow the spread of COVID-19 resulted in water demand reductions in many public, industrial, educational, and commercial buildings.^1–5^ While short periods of reduced water use (e.g., nights, weekends, school breaks) are common in building plumbing, many buildings were closed or at reduced occupancy for several months during the COVID-19 pandemic.^6–10^ Reduced water use is associated with decreased disinfectant residual levels, equilibration of hot and cold water temperatures with building temperature, and potential for increased concentrations of contaminants such as disinfection byproducts, metals, and opportunistic pathogens.^11–21^ *Legionella pneumophila*, the causative agent of Legionnaires’ disease and Pontiac fever, was of particular concern because its occurrence is widely assumed to be associated with low water use,^22–26^ and disease incidence is rising around the world.^27–30^ However, few studies have investigated its occurrence during extended periods of reduced water use.^22,26,31–33^

The quantification of *L. pneumophila* levels in potable water systems has traditionally been performed using plate culture methods. However, alternative methods such as the IDEXX Legiolert^®^ kit, a liquid culture method that yields results in most probable number (MPN) per volume, has become more widely used in recent years due to its ease of use and reported specificity for *L. pneumophila*.^34–40^ DNA-based methods (e.g., quantitative polymerase chain reaction [qPCR] and droplet digital polymerase chain reaction [ddPCR]) have also become more common due to fast turnaround times, the possibility to detect cells in the viable but non-culturable (VBNC) state, and the potential for increased specificity.^41–43^ However, comparison and interpretation of results obtained with different methods are generally lacking.^30,42^ *L. pneumophila* monitoring is usually not conducted outside healthcare settings unless the location is specifically associated with disease incidence. This lack of broad surveillance and method comparability has limited our understanding of typical *L. pneumophila* concentrations in commercial and public building plumbing, the impacts of water demand patterns in those buildings, and the effectiveness of various interventions for *L. pneumophila* mitigation.^33^

Recent investigations into the impact of COVID-19 pandemic building closures have reported that building water quality overall was negatively impacted.^7,44–49^ However, studies have reported mixed results with respect to *L. pneumophila*, including no detection of *L. pneumophila*,^44,49^ no change in occurrence as water use returned to normal levels,^45^ a small increase (2x) in *Legionella* spp. relative abundance after two months of reduced water use,^47^ and widespread detection of *L. pneumophila* that increased during closure.^46^ Each of these studies focused on individual cities, buildings, or regions, with none conducting sampling across distribution systems in different regions. Regional differences such as climate,^50^ source water, and system operation (e.g., water age, pH, residual disinfectant type) are likely to impact *L. pneumophila* occurrence in drinking water distribution systems.^35^ For example, adding a residual disinfect in drinking water distribution systems is required in most public water systems in the United States and Canada, but not in other countries, such as Switzerland.^51^

The objective of this study was to characterize the occurrence of *L. pneumophila* in drinking water collected from large, non-residential buildings at reduced occupancy due to the COVID-19 pandemic across 11 cities and three countries (United States, Canada, and Switzerland) using a culture-based method (Legiolert) and DNA-based methods (qPCR or ddPCR). Physicochemical water quality parameters, building characteristics, and details of water management practices were collected to identify factors that contribute to *L. pneumophila* occurrence. Sampling was initiated as a rapid response to assess a potential public health concern, conducted across multiple academic institutions. Site-specific investigations were conducted at several of the sampling locations, the results of which are or will be reported elsewhere.^52,53^ This study presents a cross-sectional analysis of potential *L. pneumophila* exposure risks in buildings with diverse uses, plumbing configurations, operation, and climate regions.

## 2. Methods

### 2.1 Sampling Locations

Samples were collected (n=258) from 26 buildings that were either closed or operating at reduced capacity in 11 cities in the United States, Canada, and Switzerland between April and December 2020 (Figure S1). All buildings were connected to underground municipal drinking water distribution systems and received water with free chlorine (free chlorine buildings) or monochloramine (chloramine buildings) as the secondary disinfectant, except the building in Switzerland (CH-1), which received water without a secondary disinfectant (no residual building). All buildings from each site were supplied by the same distribution system. Buildings were large, multi-story recreational, educational, office, commercial, or mixed-use (e.g., research labs and offices) facilities (Table 1). Samples were collected from showers (n=101), manual faucets (n=139), electronic (automatic) faucets (n=13), and bottle-filling stations and drinking fountains (n=5). Samples included hot (n=114), cold (n=43), or “mixed” (n=101; hot and cold water mixed prior to the fixture) water. Other relevant building information was collected from owners and maintenance staff, including building occupancy levels, building plumbing characteristics, and details of water management plans and practices prior to, during, and after building water shutdowns (Table 1 and Table S1). At Site PA, samples were collected from laboratory-scale shower rigs supplied by building water. The building plumbing at Site CH was known to contain *L. pneumophila* prior to 2020. Permission was obtained prior to sample collection, and results of testing were communicated to building owners. While the investigations at each site followed the same overall study design, there were variations in fixture flushing and analysis methods due to the collaborative nature of the sampling campaign.

**Table 1.** Summary of sampling locations and building characteristics.

| Disinfectant Type | Site | Bldg. | Source Water | Building Characteristics |  |  |  |  | Shutdown Conditions |  | Sampling Totals |  |  |
| --- | --- | --- | --- | --- | --- | --- | --- | --- | --- | --- | --- | --- | --- |
|  |  |  |  | Use Type | Number of Floors | Approx. Number of Fixtures<sup>6</sup> | Approx. Year Built (Renovation Date) | Primary Plumbing Material | Approx. Occupancy During Shutdown | Days Reduced Occupancy at Sampling | Number of Fixtures Sampled | Approx. Percent of Fixtures Sampled | Number of Samples |
| Free Chlorine | IN | IN-1 | G | Multifamily Housing | 4 | 23 | 1930 | GS | 0% | 138 - 174 | 3 | 13% | 4 |
|  |  | IN-2 |  | Commercial | 10 | 215 | 1962 | C | 0% | 120 | 4 | 2% | 4 |
|  |  | IN-3 |  | Commercial | 4 | 114 | 2002 | C | 0% | 122 | 2 | 2% | 2 |
|  |  | IN-4 |  | Mixed1 | 5 | 193 | 1951 | C | NR | 122 | 2 | 1% | 2 |
|  | OH | OH-1 | G | Education | 2 | 255 | 2012 | C | 0% | 148-191 | 4 | 2% | 4 |
|  | AZ<sup>1</sup> | AZ-1<sup>5</sup> | S & G | Mixed2 | 7 | 48 | 2018 | C | 30-50% | 162 | 7 | 15% | 7 |
|  | PA<sup>2</sup> | PA-1 | S & G | Mixed2 | 15 | 9 | 1971 (2019) | C & PVC | 5% | 125 | 9 | 100% | 18 |
|  | WV | WV-1 | S | Mixed1 | 5 | 75 | 2015 | PVC | 5% | 147 | 3 | 4% | 5 |
|  |  | WV-2 |  | Dorm | 9 | 660 | 1966 | NR | 2% | 147 | 6 | 1% | 9 |
|  |  | WV-3 |  | Dorm | 3 | 315 | 1918 | NR | 0% | 147 | 4 | 1% | 7 |
|  |  | WV-4 |  | Dorm | 10 | 550 | 1966 | C | 0% | 147 | 6 | 1% | 9 |
|  | QC | QC-1 | S | Mixed2 | 7 | 160 | 1959 (1989) | C | 2% | 62 | 21 | 13% | 21 |
|  |  | QC-2 |  | Mixed2 | 8 | 128 | 2005 | C | 2% | 53 | 12 | 9% | 12 |
|  |  | QC-3 |  | Sport complex | 5 | 290 | 1974 (2007) | C | 0% | 269 | 23 | 8% | 23 |
| Mono-chloramine | MI | MI-1 | S & G | Recreation | 3 | 95 | 1956-1978 | C & GS | 25% | 160 | 6 | 6% | 6 |
|  |  | MI-2 |  | Recreation | 2 | 37 | 1976 (2018) | C | 25% | 163 | 7 | 19% | 7 |
|  |  | MI-3 |  | Recreation | 6 | 74 | 1928 (2016) | C | 25% | 165 | 6 | 8% | 6 |
|  | VA | VA-1<sup>5</sup> | S | Mixed2 | 3 | 71 | 2005 | C | 15% | 132 | 5 | 7% | 10 |
|  |  | VA-2 |  | Mixed2 | 2 | 25 | 2007 | C | 5% | 134 | 4 | 16% | 8 |
|  | MA<sup>3</sup> | MA-1 | S | Mixed2 | 5 | 31 | 1963 | NR | 5% | 74 | 2 | 6% | 4 |
|  |  | MA-2 |  | Mixed2 | 4 | 22 | 1984 | NR | 5% | 74 | 2 | 9% | 4 |
|  |  | MA-3 |  | Mixed2 | 6 | 47 | 2017 | NR | 5% | 74 | 2 | 4% | 4 |
|  | CA | CA-1 | S | Dorm | 4 | 90 | 1990 | C | 0-4% | 106 | 8 | 9% | 8 |
|  |  | CA-2 |  | Dorm | 3 | 45 | 1940 | C | 0-2% | 106 | 6 | 13% | 6 |
|  |  | CA-3 |  | Dorm | 3 | 57 | 1950 | C | 0-2% | 106 | 6 | 11% | 6 |
| None | CH<sup>4</sup> | CH-1 | S & G | Mixed2 | 8 | >200 | 1970s | SS | <5% | 46 | 41 | ≤20% | 62 |

Sites: IN: Indiana, USA; OH: Ohio, USA; AZ: Arizona, USA; PA: Pennsylvania, USA; WV: West Virginia, USA;
QC: Québec, Canada; MI: Michigan, USA; VA: Virginia, USA; MA: Massachusetts, USA; CA: California, USA;
CH: Switzerland. NR: Not reported. Use Types: Mixed1 - Office, classroom and commercial; Mixed2 – Office and
laboratory. Source Water Types - G: groundwater, S: surface water. Primary Plumbing Materials - C: Copper, PVC:
Polyvinyl chloride, GS: Galvanized Steel, SS: Stainless Steel. <sup>1</sup>Water softening at building water inlet, varied hot
water setpoint; <sup>2</sup>Laboratory-scale shower rig, no water use during closure; <sup>3</sup>Aerator removed prior to sampling;
<sup>4</sup>Suboptimal hot water setpoint of 45°C with 2 days operated at 60°C; <sup>5</sup>green building; <sup>6</sup>Approximate number of
fixtures includes only the following potable fixtures: faucets in bathrooms and kitchens, drinking water fountains and
bottle filling stations, and showers.

### 2.2 Sample Collection

First-draw samples (n=203) were collected from every fixture sampled. Afterwards, flushed samples (n=55) were collected at a subset of locations, with flush times ranging from one to 30 minutes. Samples were collected without removing or sanitizing aerators/showerheads (except at Site MA), representing what users would experience upon opening fixtures. For both first-draw and flushed samples, 1 to 1.5 L of water was collected in a sterile container. Samples were immediately split onsite for physicochemical (100 mL), culture-based (100 mL; IDEXX Laboratories Inc., catalog number WV120SBST-20), and DNA-based (0.8 L to 1.1 L) analyses and processed the same day. Residual disinfectant was quenched using sodium thiosulfate in subsamples used for culture-based and DNA-based analyses. Sample totals by analysis method and site are provided in Table S2. Sampling controls are described in Table S3.

### 2.3 Physicochemical Analyses

Chlorine species were measured using the N,N-diethyl-p-phenylenediamine (DPD) colorimetric method and portable spectrophotometers (Hach, Loveland, CO, USA) with limits of detection (LOD) ranging from 0.02 to 0.05 mg/L as Cl_2_. Chlorine residuals are presented as total chlorine for all sites except for Site WV, where only free chlorine was measured (Table S4). Site-specific details of additional physicochemical methods (including pH and temperature) are included in Table S4. Physicochemical parameters were not recorded for samples collected at Sites CH and OH, and most samples from Site IN.

### 2.4 Culture-Based Methods

Culturable *L. pneumophila* was quantified using Legiolert according to manufacturer instructions for 100 mL potable water samples (IDEXX Laboratories, Inc., Westbrook, ME, USA, catalog number WLGT-20). The LOD is 10 MPN/L, and the upper limit of quantification (ULOQ) is 22,726 MPN/L. Positive and negative controls were included per manufacturer guidelines (Table S3).

### 2.5 DNA Concentration and Extraction

DNA was filter-concentrated onto 0.2 or 0.4 µm filters on the day of sampling (Table S5). Filters were frozen at either -80°C or -20°C until extraction. Samples were extracted using the methods described in Table S5. Filtration and extraction controls are described in Table S3.

### 2.6 qPCR and ddPCR

Of the 258 samples analyzed for culturable *L. pneumophila*, 138 were processed for *L. pneumophila* by PCR (n=120 by qPCR and n=18 by ddPCR) using previously optimized assays that were validated by the individual laboratories (referred to as Laboratories A, B, C, D, and E; Tables S6 and S7). DNA extracts from Sites CA, WV, IN, and OH were shipped overnight on dry ice to Laboratory A for qPCR (Table S7). Laboratories A, B, and E analyzed samples using qPCR with an assay targeting a single-copy macrophage infectivity potentiator (*mip*) gene.^54^ Laboratory D, which analyzed samples from Site QC, used the proprietary iQ-Check^®^ Quanti *L. pneumophila* Real-Time PCR Quantification Kit (Bio-Rad, Hercules, CA, USA, catalog number 3578103). Laboratory C analyzed samples from Site PA using ddPCR and a different mip gene assay, which was modified for use without a probe (Table S6).^55^ Additional qPCR and ddPCR details are included in Tables S6 and S7. Samples from Sites CH and MA were not analyzed for *L. pneumophila* gene targets.

All laboratories followed Minimum Information for Publication of Quantitative Real-Time PCR Experiments (MIQE) and Environmental Microbiology Minimum Information (EMMI) guidelines.^56,57^ For qPCR, all samples, standards, and controls were analyzed using triplicate reactions on each plate, except for Laboratory D, which used duplicate reactions (Table S7). Serial dilutions of synthetic DNA consisting of the 79 base pair (bp) amplicon with 30 bp neutral adaptors on each end (gBlock, Integrated DNA Technologies [IDT], Coralville, IA, USA; Table S6),^54^ ranging from 10^8^ to 10^0^ gene copies per reaction (gc/reaction) served as the standards for qPCR in all laboratories except Laboratory D, which used the proprietary standards included in the Bio-Rad kit. Efficiencies between 80 and 115% and R^2^ values of 0.98 were required for all qPCR plates containing positive samples (Table S8). Any plates that did not meet these criteria were re-run. For ddPCR, serial dilutions of synthetic DNA (gBlock, IDT)^55^ were included on each plate, as were no-template controls (Table S7). No-template controls were either negative or amplified at Cq values less than the LOD for each group.

The qPCR/ddPCR LOD and lower limit of quantification (LLOQ) were determined in each laboratory independently using serial dilutions of standards with at least 21 replicates across at least three plates, at concentrations ranging from 10^2^ to 10^0^ gc/reaction (Table S9), except for Laboratory D which followed kit instructions. The qPCR/ddPCR LOD was defined as the lowest concentration at which at least 95% of the standard replicates were detected,^56^ resulting in LODs ranging from 21 to 1×10^5^ gc/L (1 to 100 gc/reaction) across laboratories. The LLOQ was defined as the lowest concentration where the coefficient of variation was less than 25%,^58^ resulting in LLOQs ranging from 3.5×10^2^ to 2×10^5^ gc/L (6.1 to 100 gc/reaction) across laboratories. Details of the conversion of LOD and LLOQ to gene copies per liter (gc/L) are provided in Equation S1. Details of inhibition testing at each laboratory are provided in SI 3.

Cross-laboratory validation of quantitative standards was performed using a single gBlock that was sent to all laboratories using the Nazarian et al. assay.^54^ Details are provided in SI 4 and Figure S2.

### 2.7 Data Analysis

Data analyses were performed using R (version 4.1.1) and RStudio (version 1.4.1717) using a custom R pipeline that included the packages *tidyverse, lubridate*, and *readxl*.^59,60^ Because data were left-censored, imputation was performed, replacing results (physicochemical, qPCR/ddPCR, and Legiolert) less than the LOD with one half the LOD for plotting, calculating summary statistics, and non-parametric hypothesis testing. Legiolert results that were above the ULOQ were set at 30,000 MPN/L. For qPCR/ddPCR data, values between the LOD and LLOQ were replaced with the average of the LOD and LLOQ to assign identical rank for non-parametric analyses. Samples were considered positive by qPCR/ddPCR if gene copy concentrations were above the LOD. As data were non-normal (Shapiro-Wilk’s, R function *shapiro*.*test*, p<0.01), hypothesis testing was conducted using the non-parametric two-sample Wilcoxon rank sum test (“Mann-Whitney”, R function *wilcox*.*test*) with a significance threshold of 0.05. The R package *stats* was used to calculate medians. Rank correlations were calculated using Kendall’s Tau-b using *cor*.*test*, which is well-suited for nonparametric, left-censored data.^61^ Correlation analysis and principal component analysis (PCA) were performed using the R packages *vegan, Hmisc, GGally, forcats, corrplot, devtools*, and *ggbiplot*. The binomial generalized linear mixed-effects model used to investigate the relationship between *L. pneumophila* culture positivity, physicochemical parameters, and building characteristics was performed using *glmer*, with the input consisting of 112 samples from ten buildings. The equation used for model generation is provided in Figure S16. Figures were generated in R using the additional packages *ggPlot2, cowplot, scales, lubridate, repr, sf, rnaturalearth, maps, viridis, ggrepel*, and *ggnewscale*. The full R pipeline and associated data are available on GitHub (https://github.com/kathdowd/L.pneumophila_study_2022).

## 3. Results and Discussion

### 3.1 *L. pneumophila* occurrence

#### 3.1.1 *L. pneumophila* detection was building-specific

The overall sample positivity of 34% (Legiolert: n=88 of 258) to 35% (qPCR/ddPCR: n=48 of 138) suggests that *L. pneumophila* occurrence was relatively commonplace. However, detection was limited to a few specific buildings, with heterogeneity among buildings from the same distribution system. Only seven of the 26 buildings yielded Legiolert-positive samples (IN-1, AZ-1, PA-1, QC-1, QC-3, MI-3, and CH-1; Figure 1A), and the vast majority of positive samples (98%, n=86 of 88) occurred in just five buildings. Four of those five buildings were free chlorine buildings (IN-1, AZ-1, PA-1, QC-3) and the fifth was the no residual building (CH-1). In these five buildings, the percentages of Legiolert-positive samples were high (65% to 100%), and median *L. pneumophila* concentrations in Legiolert-positive samples ranged from 20 to >22,726 MPN/L. The highest concentrations of culturable *L. pneumophila* were observed in Buildings AZ-1 (median: >22,726 MPN/L) and QC-3 (median: 1,198 MPN/L), where at least one sample was >22,726 MPN/L. Similar results were observed using qPCR/ddPCR: seven buildings had at least one positive sample (AZ-1, PA-1, QC-3, WV-1, WV-2, MI-1, CA-2), three of which (AZ-1, PA-1, QC-3) were also positive by Legiolert. The median positive sample concentrations in qPCR/ddPCR-positive buildings ranged from 158 to 3.1×10^6^ gc/L, with the highest concentrations (>10^6^ gc/L) occurring in Buildings WV-1, WV-2, and QC-3 (Figure S3).

**Figure 1.**
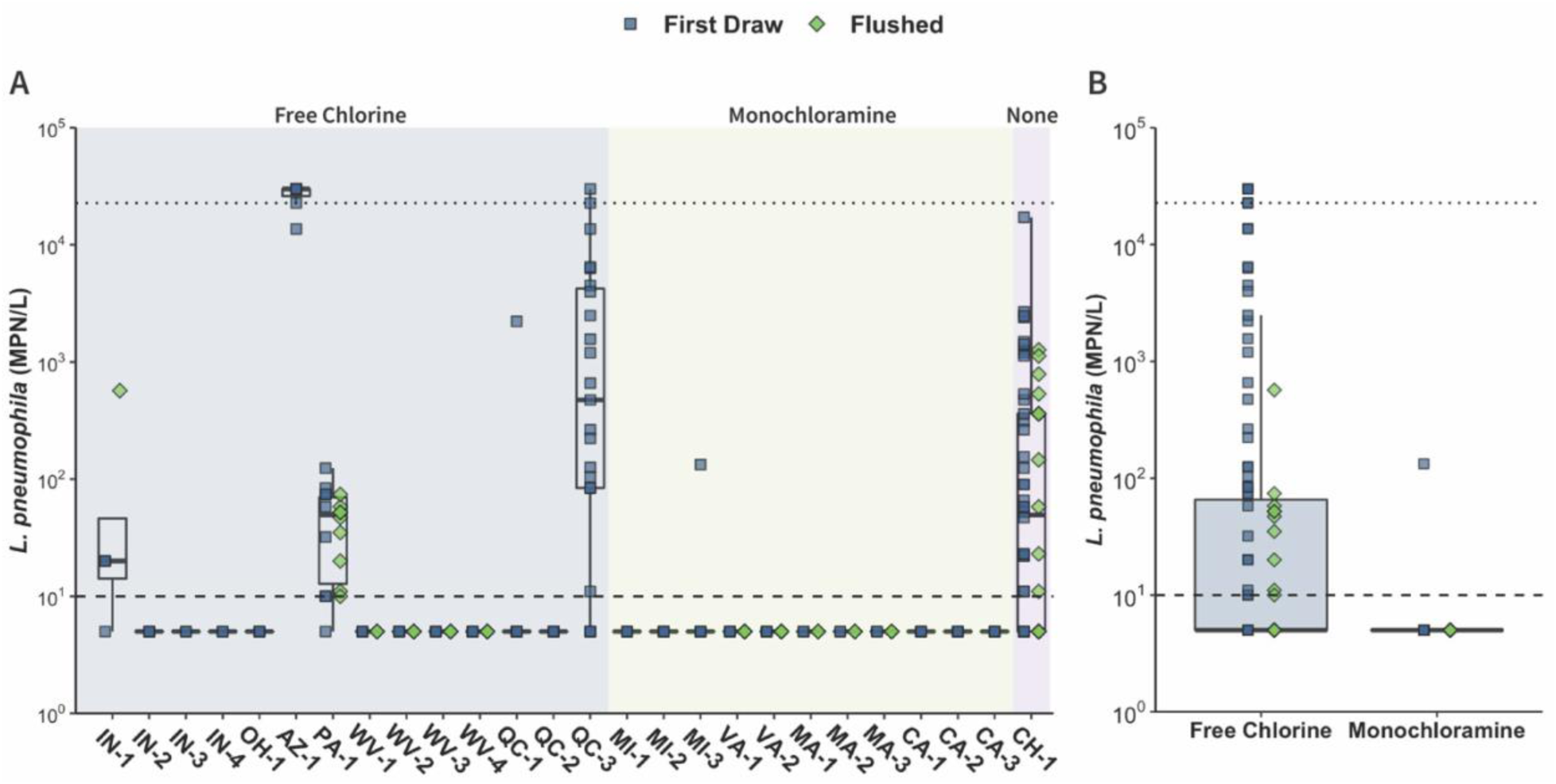
Culturable *Legionella pneumophila* concentrations (MPN/L) by (A) building and (B) secondary disinfectant type. Marker color represents flush condition, where blue circles are first-draw samples and green circles are flushed samples. The dashed horizontal line indicates the LOD (10 MPN/L) and the dotted horizontal line indicates the ULOQ (22,726 MPN/L). Results below the LOD are plotted at one-half the LOD (5 MPN/L). Results above the ULOQ are plotted as 30,000 MPN/L.

There was heterogeneity in *L. pneumophila* occurrence among different buildings in the same distribution system. For instance, at Sites IN and QC, only one building yielded multiple Legiolert-positive samples while other buildings had one or no positive samples. Buildings that were positive for *L. pneumophila* tended to have multiple positive fixtures, which has also been commonly reported in the literature.^62,63^ For example, in one recent survey, buildings with *Legionella* spp. detection in the centralized hot water systems had an average distal positivity of 83% and had significantly higher average concentrations of *Legionella* spp. compared to buildings without *Legionella* spp. in the centralized hot water system.^64^ In that same study, the majority of systems with at least one positive sample had more than 30% of distal sites positive (21 of 22 systems).

#### 3.1.2 Building-specific features shaped *L. pneumophila* positivity

The buildings with multiple Legiolert-positive samples had identifiable building plumbing operational and/or design deficiencies that did not align with industry recommended practices. Building IN-1 (Legiolert positivity: 75% [n=3 of 4]; qPCR positivity: not measured) is a large single-family home converted to a multifamily residence with an oversized water heater (100-gallon heater with 115-gallon insulated holding tank). Building AZ-1 (Legiolert positivity: 100% [n=7 of 7]; qPCR positivity: 100% [n=7 of 7]) is a green building (LEED-certified); LEED-certified buildings have previously been linked to high water residence times and, in certain cases, *L. pneumophila* occurrence.^12,65,66^ In addition, AZ-1 uses whole-building water softening upon entry of water into the building. A sample collected from a water storage tank immediately after the water softener was positive for *L. pneumophila* by both Legiolert (10,616 MPN/L) and qPCR (650 gc/L) and had a low chlorine residual (0.05 mg/L as Cl_2_), indicating that residual disinfectant loss was occurring in the water softener. Previous investigations in this building found that the softener was oversized for the building water demand, leading to high water residence time in the softener.^65,66^ Building PA-1 (Legiolert positivity: 94% [n=17 of 18]; ddPCR positivity: 67% [n=12 of 18]) is a laboratory-scale experimental shower rig consisting of three water heaters, each of which supplies three showers. The water heaters were operated at 49°C and thermostatic mixing valves limited water temperature to 40°C at the fixtures. After four months of no water use (while still maintaining the 49°C heater set point), the median *L. pneumophila* concentration in first-draw samples was 66 MPN/L. Building QC-3 (Legiolert positivity: 83% [n=19 of 23]; qPCR positivity: 91% [n=21 of 23]) is a sports building with a complex hot water system, which includes four water heaters in series. Issues identified included that a portion of the returned hot water was mixed with the hot water exiting the water heaters, resulting in a decrease of the distributed hot water temperature to below 60°C; a single thermostatic mixing valve for many showerheads (>20) created large volumes of tepid water; and the third water heater was found to be off but still connected to the system, resulting in cooling of hot water to 30°C. Building CH-1 (Legiolert positivity: 65% [n=40 of 62]; qPCR positivity: not measured) is a mixed-use building that was historically culture-positive for *L. pneumophila* and fluctuates the water heater set point between 45°C (5 days/week) to 60°C (2 days/week) to save energy. Building CH-1 also reported significant heat losses in the primary recirculating line and passive recirculation on individual floor loops, leading to suboptimal temperatures for control of *L. pneumophila*.^52^

#### 3.1.3 *L. pneumophila* occurrence varied by secondary disinfectant type

Five free chlorine buildings yielded multiple Legiolert-positive samples, whereas only a single sample was Legiolert-positive from the chloramine buildings. The positive buildings had a strong influence on overall sample positivity by secondary disinfectant residual, with samples collected from free chlorine buildings having higher Legiolert-positivity (37%, n=47 of 127) than samples collected from chloramine buildings (1.4%, n=1 of 69, Figure 1B). While samples obtained from the no residual building exhibited the highest Legiolert-positivity (65%, n=40 of 62), this building was previously reported to contain *L. pneumophila*^67^ and may not be representative of all non-chlorinated systems. Median *L. pneumophila* concentrations in the Legiolert-positive samples from the free chlorine buildings and the building with no residual were 126 and 287 MPN/L, respectively. The concentration measured in the single Legiolert-positive sample from the chloramine buildings was 133 MPN/L. Similar trends were observed in qPCR/ddPCR-positive samples, where positivity was 52% in free chlorine building samples (n=45 of 86) and 6% in chloramine building samples (n=3 of 52, Figure S4). Median concentrations in qPCR/ddPCR-positive samples were 5.3×10^3^ gc/L in the free chlorine buildings (n=45) and 1.1×10^5^ gc/L in the chloramine buildings (n=3).

In comparison to free chlorine systems, chloramine systems are commonly reported to have lower *L. pneumophila* occurrence and concentrations,^15,68–71^ even in buildings with reduced occupancy.^44^ Hypothesized reasons for lower *L. pneumophila* occurrence in chloramine systems include that monochloramine may better penetrate biofilms,^72–75^ may provide more persistent residuals in building plumbing,^76^ and may more efficiently inactivate amoebal hosts or *L. pneumophila* within amoebae.^77,78^ The few studies that examined *L. pneumophila* in distribution systems without a disinfectant residual generally reported low prevalence of *L. pneumophila*,^55,79–82^ though there have been reports of *L. pneumophila* occurrence in some building hot water systems.^83,84^

### 3.2 Impacts of physicochemical parameters and flushing

#### 3.2.1 Secondary disinfectant concentration influenced presence and concentrations of *L. pneumophila*

Despite reduced occupancies in the buildings and the limited use of water quality mitigation measures (Table 1 and Table S1), 69% (n=101 of 146) of the first-draw samples analyzed for chlorine contained a chlorine residual at or above the detection limit (≥0.02 or ≥0.05 mg/L as Cl_2_). This relatively high level of chlorine positivity in samples may be linked to recent use of fixtures, as access to study fixtures was not restricted at all sites prior to sampling. Median chlorine residuals were lower in first-draw samples compared to flushed samples in both free chlorine (first-draw: <0.05 mg/L as Cl_2_, flushed: 0.05 mg/L as Cl_2_) and chloramine (first-draw: 0.13 mg/L as Cl_2_, flushed: 0.88 mg/L as Cl_2_) buildings (Figure S5A). First-draw samples with measurable chlorine residuals were not limited to buildings with higher occupancies and those conducting preventative maintenance, but were also collected from reduced occupancy buildings (≤5%). Even with few occupants, some buildings may have some water demand due to leaks, maintenance activities, air conditioning systems or cooling towers, and operation of treatment systems (e.g., water softeners that automatically regenerate). In contrast, other investigations of COVID-19 related building closures have reported that measurable disinfectant residuals were lacking in most samples.^44,48^

Overall, *L. pneumophila* positivity in samples tended to decrease with increasing disinfectant residual (Figures 2A, S6, and S7). In free chlorine buildings, approximately 80% (78%, n=35 of 45 by Legiolert; 82%, n=37 of 45 by qPCR/ddPCR) of positive samples occurred when the chlorine residual was less than or equal to 0.1 mg/L as Cl_2_ (Figures S6C and S6D). Only three free chlorine samples were Legiolert-positive with chlorine residuals above 0.2 mg/L as Cl_2_, and the highest chlorine residual in the Legiolert-positive samples was 0.4 mg/L as Cl_2_. The single Legiolert-positive sample from a chloramine building had a chlorine residual below the detection limit. However, the other 27 chloramine samples that contained little to no chlorine residual (≤0.1 mg/L as Cl_2_) were Legiolert-negative.

**Figure 2.**
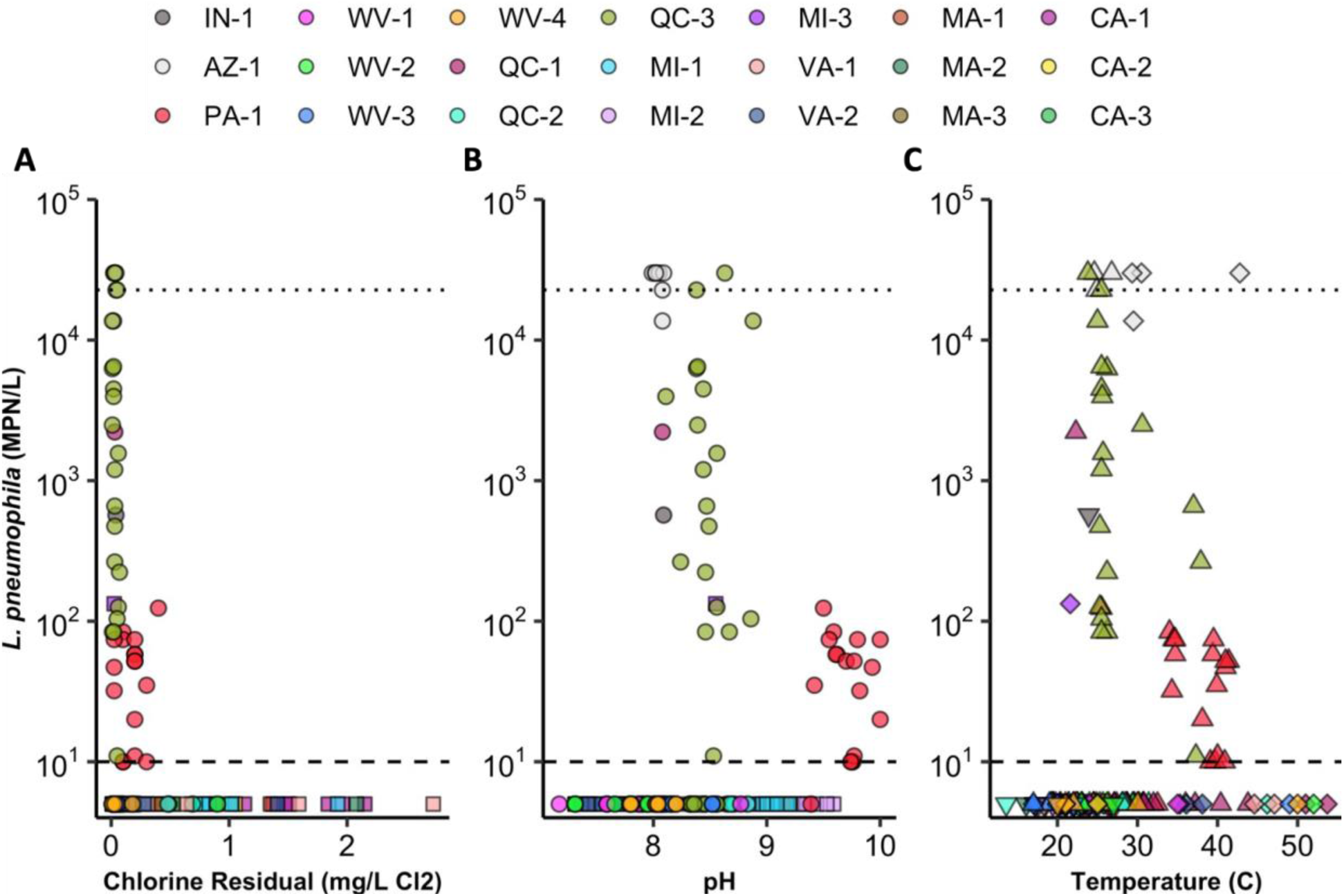
Physicochemical parameters and culturable *L. pneumophila* (MPN/L) for samples by building. A) chlorine residual (mg/L as Cl_2_), B) pH, and C) temperature. For A and B, marker shape represents secondary disinfectant type, with circles being free chlorine and squares being monochloramine. For C, marker shape shows the water temperature type, with upside down triangles being cold water samples, diamonds being hot water samples, and triangles being hot and cold water mixed. The dashed line shows the LOD. The dotted line shows the ULOQ. Buildings CH-1, IN-2, IN-3, IN-4, and OH-1 are excluded because physicochemical parameters were not recorded.

Free chlorine concentrations of at least 0.2 mg/L as Cl_2_ have been recommended for effective control of *Legionella* spp. in building plumbing.^30,85^ In a survey of residences, a free chlorine residual less than 0.4 mg/L as Cl_2_ at the cold water inlet was associated with significantly higher *Legionella* spp. gene copy numbers and household positivity.^86^ However, another study reported no relationship between incoming free chlorine concentration and *Legionella* spp. occurrence.^64^ While higher concentrations of monochloramine have been found to decrease the occurrence of *L. pneumophila*, the concentration required for prevention of *L. pneumophila* varies in the literature._30,85,87–90_

#### 3.2.2 Sample temperature and pH were not associated with *L. pneumophila* culture-positivity

Of the samples where temperature was measured (n=182), first-draw (n=146) water temperatures were similar to ambient building temperatures regardless of their source (hot, cold, or mixed), with an overall median of 25°C (Figure S5C). Only 19% (n=4 of 21) of cold water first-draw samples were below 20°C (median: 23°C), and only 5% (n=2 of 39) of hot water first-draw samples were above 50°C (median: 22°C, Figure 2C). In paired first-draw and flushed samples where temperature was recorded (n=28 pairs), flushing significantly changed the water temperature, reducing the cold water temperatures and increasing hot and mixed water temperatures (p<0.05, Figure S8).

Temperatures in Legiolert-positive hot (n=5) and mixed (n=40) samples ranged from 22°C to 43°C, which is within the suitable growth range for *L. pneumophila* (20°C to 45°C),^63,91,92^ and did not show a trend with *L. pneumophila* occurrence or concentration (Figure 2C). Culture-negative samples occurred at all temperatures measured (n=136, 14°C to 54°C) and sample temperature types (hot, cold, and mixed). The majority of Legiolert-negative samples (82%, n=111 of 136) were also at temperatures within the *L. pneumophila* suitable growth range. In samples analyzed using qPCR/ddPCR for which temperature was recorded (n=130), the temperature in the positive samples (n=48) ranged from 20°C to 43°C, while in negative samples (n=82) temperatures ranged from 17°C to 52°C (Figure S9). The highest concentrations of *L. pneumophila* (>1×10^6^ gc/L) detected by qPCR/ddPCR occurred in two hot water samples from Buildings WV-1 and WV-2 and one mixed water sample from Building QC-3, which were at approximately room temperature (20 to 26°C).

It is recommended that hot water temperatures be at least 60°C in water heaters and 55°C at distal points to control *L. pneumophila*.^30,93^ However, observed water temperature at distal fixtures may not always be helpful in predicting *L. pneumophila* concentrations because distal sampling locations acclimate to ambient temperature.^64,86,94^ Therefore, approaches that incorporate whole-building temperature profiling may be more effective for assessing the influence of hot water temperature on *L. pneumophila*.^91^ Intermittent heating, and other energy-saving methods have previously been linked to lower water qualities due to decreased hot water temperatures in building plumbing.^95,96^ Consistent with this, two buildings with more than one culture-positive sample (CH-1 and AZ-1) practiced hot water temperature fluctuation for energy savings.

Additional physicochemical parameters, including pH (Figures 2B and S10), dissolved oxygen (Figure S11), and electrical conductivity (Figure S12) were measured in a subset of samples (Tables S4 and S10). Sample pH in chloramine buildings (median: 9.1, n=69) was higher than the pH of samples from free chlorine buildings (median: 8.2, n=112; Figures 2B and S5B) most likely because higher pH is a strategy to reduce nitrification. pH values in Legiolert-positive samples ranged from 8.0 to 10.0. Though the median pH was higher in Legiolert-positive samples (8.6, n=46) than in Legiolert-negative samples (8.3, n=135), pH tended to co-vary by building. In paired samples where pH was measured (n=27), pH was significantly different between first-draw and flushed samples (p<0.05).

#### 3.2.3 Fixture flushing did not consistently reduce *L. pneumophila* occurrence

The impact of flushing during sampling on *L. pneumophila* was investigated using paired samples from 48 fixtures (n=96 samples from 12 buildings). While flushing (for two to 30 minutes, median: five minutes) tended to increase chlorine residual (Figure S13) and shift water temperatures toward hot/cold water set points (Figure S8), the impact of flushing on Legiolert-positivity varied by fixture: 24 pairs were Legiolert-negative in both first-draw and flushed sample, 18 were positive in both, three fixtures went from negative to positive (PA-1 and CH-1), and three fixtures went from positive to negative with flushing (CH-1; Figure S14). With qPCR/ddPCR, nine fixtures were negative in first-draw and flushed samples, three fixtures were positive in both samples (PA-1), four fixtures went from negative to positive (PA-1 and WV-1), and four went from positive to negative (in PA-1 and WV-2; Figure S15). When only considering Legiolert-positive pairs (n=18 pairs from three buildings [PA-1, CH-1, and IN-1]) flushing did not significantly change the concentration of culturable *L. pneumophila* (median decrease of 27 MPN/L with flushing, p=0.24). However, at individual sampling locations, flushing increased *L. pneumophila* concentrations as much as 549 MPN/L (IN-1) and decreased concentration by as much as 1.6×10^4^ MPN/L (CH-1). In the three qPCR/ddPCR-positive sample pairs (PA-1), the impact of flushing on *L. pneumophila* gene copies was similarly mixed (*L. pneumophila* concentrations increased in one fixture and decreased in two). Therefore, flushing associated with sampling showed mixed results for remediation of *L. pneumophila* in study buildings.

Most guidance recommends volume- or water-quality-based flushing of the full building water system, which was not evaluated in this study.^97–99^ Other studies have also reported that flushing that is not performed systematically or that does not consider building plumbing characteristics may be ineffective for improving building water quality.^100,101^ While water management practices varied by building, only one (OH-1, of n=21 respondents) reported having a formal building water management plan that predated the pandemic and one (CH-1) reported a pre-existing informal management plan. Only two buildings (MI-1 and MI-2) of the 17 buildings for which additional information was provided reported regularly flushing building plumbing while occupancy was restricted (Table S1). MI-1 tended to have higher first-draw disinfectant residuals (median: 0.8 mg/L as Cl_2_, n=6) than the disinfectant residuals in first-draw samples from other chloramine buildings (median: 0.09 mg/L as Cl_2_, n=48). None of the samples from MI-2 (n=7) contained measurable concentrations of disinfectant, but fixtures in this building were restricted from use for one week prior to sampling. Interestingly, samples from MI-1 were among the only samples from chloramine buildings where *L. pneumophila* was detected by qPCR, though the results were below the LLOQ (Figure S3).

#### 3.2.4 Fixture type was not associated with differences in *L. pneumophila* occurrence

Showers and other fixtures that generate droplets and aerosols are of particular concern for opportunistic pathogen respiratory infections because inhalation is the primary route of exposure. ^102–104^ Among showers and manual faucets, the primary fixture types in this study, overall culture-positivity was similar (showers: 38%, n=38 of 101; manual faucets: 34%, n=47 of 139). By qPCR/ddPCR, showers were 45% positive (n=38 of 84) and manual faucets were 11% positive (n=4 of 36). Positivity in electronic faucets (n=13 samples across four buildings), which have been linked to *Legionella* spp. occurrence,^12,14,95,105,106^ was 23% by Legiolert (n=3, AZ-1) and 46% by qPCR (n=6, AZ-1 and WV-1).

#### 3.2.5 Combined effects of quantitative physicochemical and building parameters in *L. pneumophila* occurrence

To investigate the impact of building characteristics and physicochemical parameters on *L. pneumophila* culture-positivity in free chlorine buildings, a binomial generalized linear mixed model (glmm) was developed. The model was developed using free-chlorine samples from 10 buildings where associated physicochemical measurements and building characteristics were available (n=112) because disinfectant type has been previously shown to influence *L. pneumophila* occurrence.^68,69,107^ The model included the physicochemical parameters pH, temperature, and free chlorine concentration and the building-specific parameter building age as fixed effects; these parameters were chosen based on measured factors previously reported to influence *L. pneumophila* occurrence.^62,80,108–110^ To account for clustering by building, building identity was included in the model as a random effect. This analysis showed that none of the parameters were significantly associated with *L. pneumophila* positivity (p>0.05, Figure S16). Though no parameters met the significance threshold, pH was the model input with the greatest effect on the probability of *L. pneumophila*-positivity (p=0.14, odds ratio: 10.9; see Figure S16 for equation). A principal component analysis (PCA) was also used to investigate the impact of multiple parameters on *L. pneumophila* occurrence. *L. pneumophila* positivity again could not be fully explained by the quantitative variables included, but most Legiolert-positive samples clustered in a region representing lower chlorine concentrations (Figure S17).

### 3.3 *L. pneumophila* quantification with Legiolert versus qPCR/ddPCR

*L. pneumophila* was quantified in samples from 17 buildings using qPCR (n=120) and one building using ddPCR (PA-1; n=18). Of these samples, 35% (n=48 of 138) were positive by qPCR/ddPCR and 34% (n=88 of 258) were positive using Legiolert, with an 87% (n=120 of 138) agreement between qPCR/ddPCR and Legiolert (positive by both methods: n=37; negative by both methods: n=83). The 13% (n=18 of 138) of samples where there was disagreement between the two methods included 11 samples that were positive only by qPCR/ddPCR (PA-1, QC-3, WV-1, WV-2, MI-1, CA-2) and seven samples that were positive only by Legiolert (PA-1 and MI-3). The samples that were only positive by Legiolert exhibited low concentrations (median: 32 MPN/L), and all but one (n=6 of 7) were collected from PA-1 and analyzed using ddPCR, indicating that the ddPCR LOD and/or DNA extraction recovery efficiency may have impeded detection. However, false positives using Legiolert have also been reported (3% to 4%),^111–113^ which could have contributed to this discrepancy. The majority of samples that were positive only by qPCR/ddPCR (n=9 of 11) were quantifiable (above the LLOQ), spanning the full range of *L. pneumophila* concentrations (10^2^ to 10^6^ gc/L, Figure 3A), indicating the presence of dead or non-culturable *L. pneumophila*.

**Figure 3.**
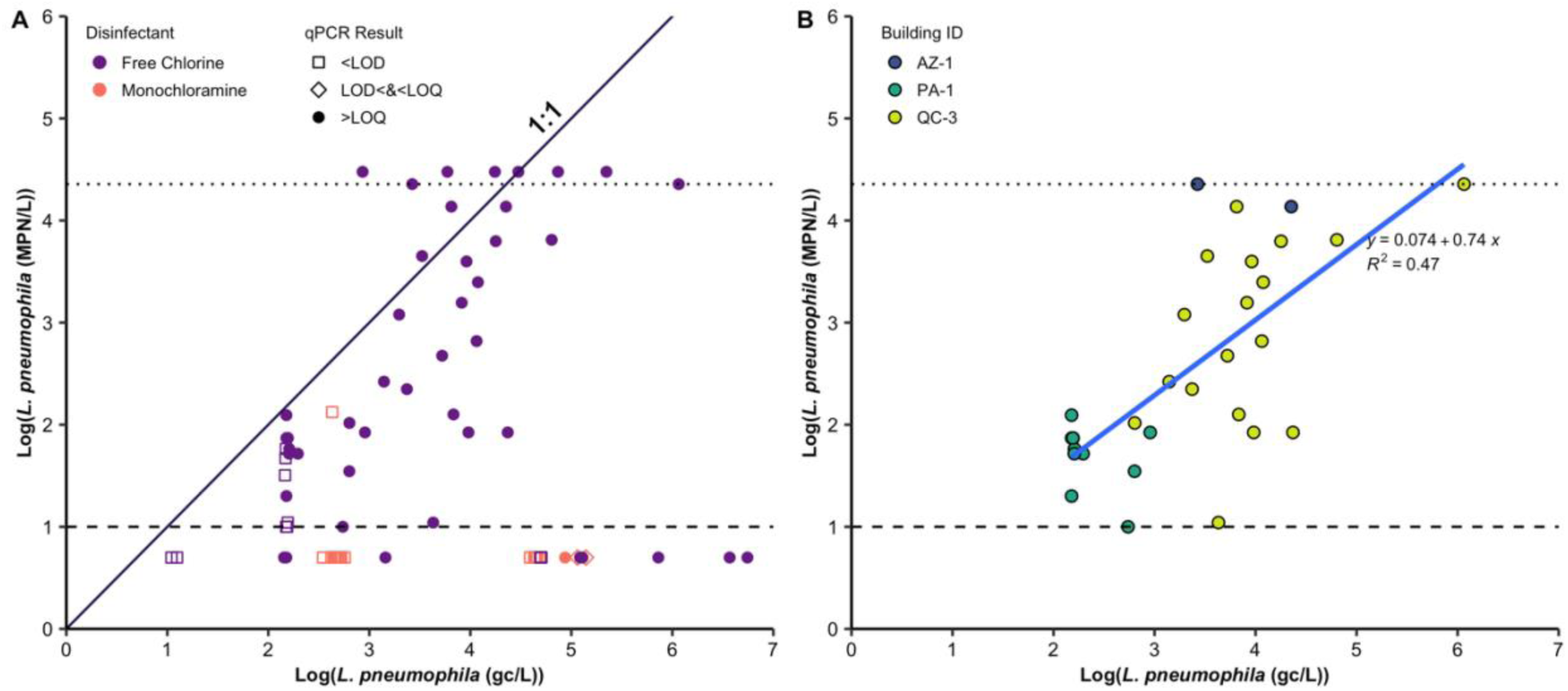
Culturable *L. pneumophila* results (log(MPN/L)) versus qPCR/ddPCR results (log(gc/L)). A) Results for all samples analyzed by both Legiolert and qPCR/ddPCR (n=138). The solid black diagonal line has a slope of 1 for reference. Samples that were below the LOD were plotted at one half the LOD (open squares). qPCR/ddPCR samples between the LOD and LLOQ were plotted as the average of the LOD and LLOQ (open diamonds). Samples above the LLOQ were plotted as filled circles. Results above the Legiolert ULOQ were plotted at 30,000 MPN/L. B) Results for the subset of samples that were quantifiable by both methods (n=31). The blue line is the linear regression. The dashed horizontal line indicates the Legiolert LOD (10 MPN/L) and the dotted horizontal line indicates the Legiolert ULOQ (22,726 MPN/L).

Samples positive and quantifiable by both methods had a median 0.72 log more *L. pneumophila* measured by qPCR/ddPCR compared to Legiolert (n=31, Figure 3B). However, seven samples that were positive by both methods yielded higher results with Legiolert than qPCR/ddPCR (median difference: 0.32 log). Linear regression analysis of the quantifiable samples resulted in a line with a slope of 0.74 and y-intercept of 0.07 (R^2^=0.47, Figure 3B). This analysis was used to estimate the relationship between qPCR/ddPCR and Legiolert, but, as shown in Figure 3B1, this relationship may be influenced by location-specific characteristics. Among all samples analyzed by qPCR and Legiolert, there was a negative correlation (Kendall’s tau=-0.277, p<0.001, n=138), likely due to the way non-quantifiable values were substituted. When only the quantifiable data are considered, the correlation is stronger and positive (Kendall’s tau=0.493, p<0.001, n=31).

Previous studies have reported higher *Legionella* percent positivity by qPCR than culture-based methods, attributing this difference to higher sensitivity of DNA-based methods and the inclusion of cells that are non-viable and/or non-culturable.^114–116^ However, the relatively high LODs of some of the laboratories (100 gc/reaction, translating to up to 10^5^ gc/L; Table S9), as well as losses during concentration and extraction, may have prevented the detection of low concentrations of *L. pneumophila* DNA in some samples. The higher concentrations observed with qPCR/ddPCR compared to Legiolert agree with previous studies, which have reported that qPCR results were 0.5 to 3 log higher than the corresponding culture-based results.^42,85,115^ The correlations between quantifiable samples (Kendall’s tau=0.493; linear regression slope=0.74; Figure 3B) were similar to those reported in previous studies comparing qPCR to culture-based methods.^116,117^

## 4. Broader Implications

### 4.1 Implications for building water management

The proper design and management of building plumbing is critical for *L. pneumophila* control.^30^ Most existing guidance on *L. pneumophila* control in building plumbing advocates for the development of formal building water management plans or programs.^30,97,118^ As with the buildings in this study, few non-healthcare building water systems have a program due to lack of awareness, expertise, and resources.^119^ Furthermore, even when a management program and knowledge of best practices exist, they are not always executed properly.^69,119^ This study demonstrates that assessing the engineering and water quality aspects of the building plumbing can identify possible causes of *L. pneumophila* occurrence without having to fully implement a water management program.

Most guidance on *L. pneumophila* prevention and building recommissioning recommend flushing to introduce fresh water from the distribution system into building plumbing.^98,99,118,120,121^ The mixed effectiveness of flushing in this study emphasizes the importance of developing building-wide *L. pneumophila* management plans that consider building-specific plumbing characteristics.^3,122^ Overall, there is a need for the development of evidence-based guidance for *L. pneumophila* prevention and remediation in building plumbing that is accessible to broad audiences and can be implemented affordably.^123,124^

### 4.2 Implications for monitoring *L. pneumophila* in non-healthcare buildings

As there are few resources available to aid practitioners in the selection of *L. pneumophila* monitoring methods, it can be challenging to determine which methods are best-suited to a particular application. Action levels for *L. pneumophila* are primarily based on plate culture methods, such as ISO 11731, which yield results in colony forming units (CFU) per volume.^30^ However, liquid culture and DNA-based methods like those used in this study are increasingly being used for environmental monitoring due to their ease of use and reduced time to results.^41–43^ Additional studies that compare the performance of these methods in environmental matrices and realistic building management scenarios and relate the results to health outcomes and risk assessment are needed to bridge the gap between results and action limits.

Due to the variation in existing guidance and action limits, it is difficult to determine if remediation (e.g., shock chlorination) is required when *L. pneumophila* is detected in building plumbing. *L. pneumophila* action levels that have been proposed by various publications and organizations vary widely: risk-based limits vary from 14.4 CFU/L (disability adjusted life year, 10^−6^ DALY target) to 1,410 CFU/L (10^−4^ annual probability of infection target) in non-healthcare showers and from 12.3 CFU/L to 4,670 CFU/L for total building water fixture exposures;^125^ 1,000 CFU/L is recommended by the European Working Group for *Legionella* (EWGLI);^126^ and health outcome-based levels are as high as 50,000 CFU/L.^30^ To place MPN and gene copy results in a health context, they must be converted to approximate CFU. Previous studies have found MPN to be equivalent to approximately 1.2 times CFU concentrations.^111,127^ Using this approximation, the number of samples in the current study that would exceed action thresholds ranged from 33 samples from five buildings using the adjusted Hamilton et al. 14.4 CFU/L threshold,^125^ 25 samples from four buildings if using the adjusted EWGLI 1,000 CFU/L threshold,^126^ and up to three samples (samples above the Legiolert ULOQ) when using the adjusted 50,000 CFU/L threshold.^30^ As demonstrated by this variation, there is a need for greater clarity on which *L. pneumophila* concentrations should trigger mitigation, and recommendations should consider the vulnerability of the population exposed.^26,30^

### 4.3 Implications for communicating health risks to the building community and the general public

Based on disease surveillance data, available evidence does not support that periods of reduced water demand led to higher incidence of legionellosis. Overall reporting of legionellosis cases was lower in the United States in 2020 than in recent years prior to the pandemic,^128^ though some regions reported similar or higher case numbers in 2020.^129,130^ Switzerland also reported fewer cases in 2020 than expected based on recent increasing trends,^131^ and an increase in cases was not observed after travel and work-from-home restrictions associated with the first wave of COVID-19 were lifted.^132^ Diagnosis of legionellosis may have decreased in some areas during 2020 due to the overburdening of healthcare systems and the potential for undiagnosed co-infections with SARS-CoV-2, though the occurrence of *L. pneumophila* and SARS-CoV-2 co-infections has only been reported in one of three studies investigating this topic.^133–135^

Significant attention was paid to the potential public health threat posed by exposure to *L. pneumophila* in building water systems during pandemic building closures. This study, which included data from geographically and structurally diverse buildings in three countries, found that approximately 20% of buildings contained *L. pneumophila* concentrations in showers that were potentially of health concern (using the Hamilton et. al 14.4 CFU/L threshold for showers^125^ multiplied by 1.2 to convert to approximate MPN/L), suggesting that there was not widespread occurrence of *L. pneumophila* in buildings with reduced occupancy. Limitations of this study include that few sites were monitored for *L. pneumophila* prior to COVID-19 pandemic building closures, which prevents investigation of how *L. pneumophila* concentrations changed over time with reduced water use in buildings, and that detailed water use data were not available at most sites. In addition, it is important to note that distal site positivity does not necessarily predict incidence of Legionnaires disease, which also depends on exposure route and population susceptibility.^136,137^ Therefore, even the moderate overall positivity and high positivity in some individual buildings in this study do not necessarily indicate high probability of disease incidence, and no legionellosis outbreaks were associated with buildings in this study. However, due to lack of data availability, it is unknown to what degree the plumbing and water quality characteristics of the buildings sampled are representative of those within and across countries. Additionally, other water quality issues that can develop during prolonged periods of stagnation, such as occurrence of other opportunistic pathogens and high concentrations of metals, were not assessed in this work and are similarly not well studied. While sites receiving monochloramine-treated water were overwhelmingly negative for *L. pneumophila*, it is important to recognize limitations and trade-offs to this observation. Most buildings cannot choose which residual disinfectant is supplied in the water, and there are other concerns with monochloramine that should be considered when evaluating secondary disinfectants such as the potential for nitrification, formation of unregulated disinfection byproducts, and growth of other opportunistic pathogens.^19,138,139^ Additional work is needed to fully characterize the impact of reduced water use periods on building water quality and inform how buildings can prevent or react to reduced water use.

## Data Availability

All data produced are available online at https://github.com/kathdowd/L.pneumophila_study_2022

## Acknowledgements

The authors would like to thank building owners and managers that responded to questions and provided facility access and acknowledge IDEXX Laboratories, Inc. for providing free and discounted Legiolert kits. The authors appreciate Aliya Ehde, Kirk Olsen, Benjamin Gincley, Umang Chauhan, Irmarie Cotto, and Katherine Vilardi for their water sampling assistance and Amy Pruden, Marc Edwards, and Frederik Hammes for their review of the manuscript. K.S.D. was supported by the National Science Foundation (NSF) Graduate Research Fellowship under Grant No. DGE-1256260, the University of Michigan Blue Sky Grant, and the University of Michigan Rackham Predoctoral Fellowship. H.D.G. was supported by the National Science Foundation (NSF) Graduate Research Fellowship under Grant No. DGE-1752814. S.M.J was supported by Arizona State University engineering startup funds. S.P was supported by a University of Pittsburgh Central Development Research award. W.J.R. was supported by EAWAG discretionary funding. C.R.P., A.J.W., and C.L. were supported by NSF Grants No. CBET-2039498 and CBET-2027049. C.R.P. was supported by the Purdue University Lilian Gilbreth Postdoctoral Fellowship. S.V. and L.H. were supported by NSF Grants No. CBET-2029850 and CBET-1749530. Water Research Foundation Project 4721 additionally supported the development of qPCR/ddPCR methods applied in this study.

