## Supplemental Information (SI) for "*Legionella pneumophila* occurrence in reduced-occupancy buildings in 11 cities during the COVID-19 pandemic"

* Corresponding authors
** Co-first authors

**Contents**

1. SI 1 – Methods Supplement…………………………………………………….........S5
    a. SI 1.1 – Sampling Locations………………………………………………… S5
    b. SI 1.2 – Sampling and Analysis……………………………………………… S8
2. SI 2 – qPCR and ddPCR Quality Assurance and Quality Control …………………… S15
3. SI 3 – qPCR and ddPCR Inhibition Testing Summary……………………………….. S18
4. SI 4 – qPCR and ddPCR Cross-Laboratory Validation Testing……………………… S20
5. SI 5 – Results Supplement………………………………………………………… S21

**List of Figures**

1. **Figure S1.** Sampling site geographic locations in the U.S., Canada, and Switzerland as well as distribution system disinfectant type.
2. **Figure S2.** Cross-laboratory validation results for the laboratories using the Nazarian et al. (2008) qPCR assay.
3. **Figure S3** *L. pneumophila* qPCR/ddPCR results by building and secondary disinfectant type.
4. **Figure S4**. *L. pneumophila* qPCR/ddPCR results by disinfectant type and sample type.
5. **Figure S5.** A) Chlorine residual, B) pH, and c) temperature by building and sample type.

6. **Figure S6.** *L. pneumophila* Legiolert and qPCR/ddPCR results by disinfectant type and sample type.
7. **Figure S7.** *L. pneumophila* qPCR/ddPCR results as a function of sample chlorine residual.
8. **Figure S8.** Sample temperature results by condition (first-draw vs. flushed) and fixture temperature (cold, hot, or mixed) for paired samples.
9. **Figure S9.** *L. pneumophila* qPCR/ddPCR results as a function of sample temperature.
10. **Figure S10.** *L. pneumophila* qPCR/ddPCR results as a function of sample pH.
11. **Figure S11.** Dissolved oxygen as a function of culturable *L. pneumophila*.
12. **Figure S12.** Electrical conductivity as a function of culturable *L. pneumophila*.
13. **Figure S13.** Sample chlorine residual results by condition (first-draw vs. flushed) and disinfectant type (free chlorine and chloramine) for paired samples.
14. **Figure S14.** *L. pneumophila* Legiolert results (MPN/L) by condition (first-draw vs. flushed) for paired samples.
15. **Figure S15.** *L. pneumophila* qPCR/ddPCR results (gc/L) by site for paired samples.
16. **Figure S16.** Generalized linear mixed effects model input and results for free chlorine samples with associated physicochemical measurements and building characteristics.
17. **Figure S17.** Principal component analysis (PCA) incorporating physicochemical parameters (chlorine residual, temperature, and pH) and building characteristics (number of floors and building age)

**List of Tables**


1. **Table S1.** Additional buildings, sampling, occupancy, and preventative measure information.
2. **Table S2.** Summary of total number of samples totals by site, flush condition, and type of analysis.
3. **Table S3.** Summary of sampling and analysis controls
4. **Table S4.** Summary of physicochemical methods.
5. **Table S5.** DNA collection, extraction, and quantification.
6. **Table S6.** qPCR and ddPCR primers, probes, and standards.
7. **Table S7.** qPCR and ddPCR protocols.
8. **Table S8.** qPCR/ddPCR standard curve parameters.
9. **Table S9.** qPCR/ddPCR limit of detection (LOD) and lower limit of quantification (LLOQ) testing results in gene copies per reaction (gc/rxn).
10. **Table S10.** Physicochemical results summary.

### SI 1 – Methods Supplement

#### SI 1.1 – Sampling Locations

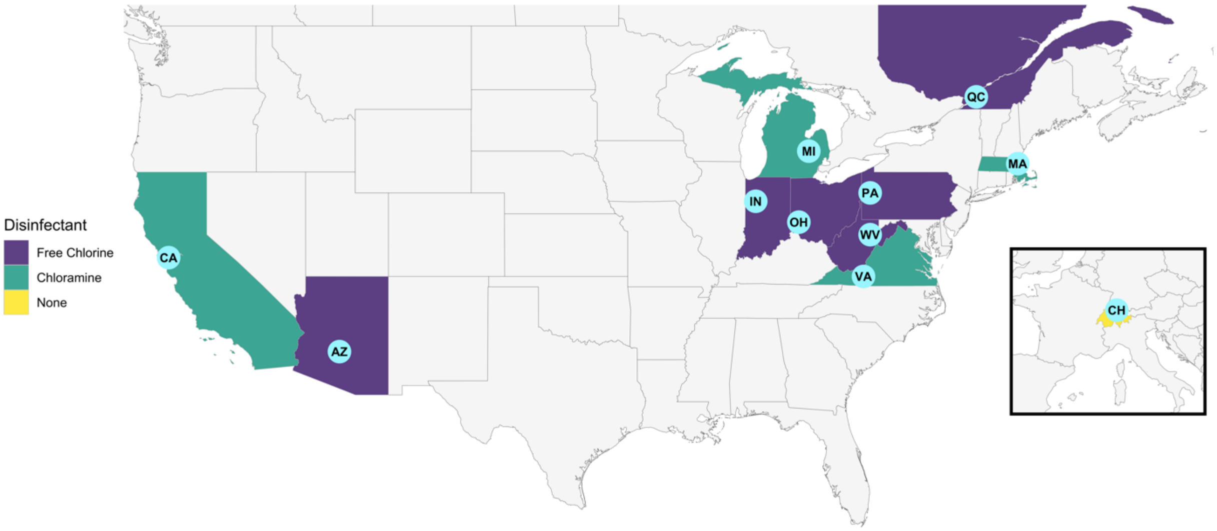


**Figure S1.** Sampling site geographic locations in the U.S., Canada, and Switzerland as well as distribution system disinfectant type. Sampling site fill color represents the type of secondary disinfectant (free chlorine, chloramine, or no residual disinfectant) used in the distribution system serving the buildings sampled.

**Table S1.** Additional buildings, sampling, occupancy, and preventative measure information.

| Site | Building | State or Region | Disinfect. | Type | Hot Water Recirc. | Closure Date | Sampling Date(s) | Description of Occupancy/Closures | Preventative Measures During Low Occupancy Period | Measures after Study, Follow-Up, or Response Actions |
|---|---|---|---|---|---|---|---|---|---|---|
| IN | IN-1 | Indiana | Free Chlorine | Full-Scale | Yes | 3/23/20 | 8/5/20, 8/12/20 | Completely unoccupied. | No preventative measures were taken prior to sampling. | Flushing was performed after *L. pneumophila* was detected by Legiolert. Follow-up sampling was then conducted. |
| | IN-2 | | | | Yes | 3/23/20 | 7/21/20 | Mostly unoccupied. | | Some flushing was conducted by the building operators. |
| | IN-3 | | | | Yes | 3/23/20 | 7/23/20 | | | |
| | IN-4 | | | | Yes | 3/23/20 | 7/23/20 | | | |
| OH | OH-1 | Ohio | Free Chlorine | Full-Scale | Yes | 3/15/20 | 8/15/20 | Building was closed from March - September 2020 | The utility operator flushed the building in early August to attempt to get a consistent chlorine residual. | Flushing and shock chlorination performed after *L. pneumophila* detection. |
| AZ | AZ-1 | Arizona | Free Chlorine | Full-Scale | Yes | 3/15/20 | 8/24/20 | At the lowest occupancy during COVID-19 pandemic (March 2020), occupancy was reduced to 15-25% of normal occupancy. By Aug-Sept 2020 during the sampling period, occupancy rose to 30-50% and 40-70% respectively, depending on the floor considered (1-5). | No preventative measures prior to *L. pneumophila* detection by this study. Some previous studies on physical chemical water quality indicated other issues in the building such as lack of chlorine residual (potential removal by water softener) and DBP formation, however no actions taken until to *L. pneumophila* detection. | Building was flushed after to *L. pneumophila* detection. Shower was flushed for 5 h by the facilities manager. 9/4/20 sinks, janitor's closets, and showers were flushed for 30 mins on every floor in stages of even/odd floors. Water heater set point was turned to 140F and allowed to recirculate 30-35 mins then returned to 115 degrees. 9/8/20 janitors did periodic (unspecified) flushing. Resin tanks regenerated on a weekly basis. |
| PA | PA-1 | Pennsylvania | Free Chlorine | Lab-Scale | No | 3/19/20 | 7/22/20 | Shower rig was completely stagnant prior to sampling. Building was significantly stagnant until early June when partial re-opening began. | No preventative measures were taken prior to sampling. | None |
| WV | WV-1 | West Virginia | Free Chlorine | Full-Scale | Yes | 3/13/20 | 8/7/20 | Occupancy reduced to ~5% as of 3/13/20 | No preventative measures were taken prior to sampling. | None |
| | WV-2 | | | | Yes | 3/13/20 | 8/7/20 | Occupancy reduced to ~2% as of 3/13/20 | | |
| | WV-3 | | | | Yes | 3/13/20 | 8/7/20 | Unoccupied except for occasional maintenance. | | |
| | WV-4 | | | | Yes | 3/13/20 | 8/7/20 | | | |
| QC | QC-1 | Quebec (CA) | Free Chlorine | Full-Scale | Yes | 3/13/20 | 5/14/20 | Occupancy reduced to approximately 2% as of 3/13/20, increased to approximately 5% over the summer. | Building water was being used by the HVAC system but was not intentionally flushed or managed. | Full recommissioning flushing following Quebec's procedures performed on 5/8/20. Building engineers designed a flushing plan for all water points. Building has a newly developed flushing plan. |
| | QC-2 | | | | Yes | 3/13/20 | 5/5/2020 | | | |
| | QC-3 | | | | Yes | 3/13/20 | 12/7/20 | Approx. < 5%; no visitors as of 3/13/20 (only maintenance and managers). Day camps as of July 1st, 2020 (no shower use, but increased occupancy) | Building water was being used by the HVAC system but was not intentionally flushed or managed. Partial recommissioning flushing (only showers were flushed for 5-min, mitigated water) on 7/14/20. Showers remained closed till now due to elevated *L. pneumophila* concentrations. | Building was flushed (only showers, mitigated water, 5-min) on 7/14/20. Showers remained close till 2021, at least. |

| Site | Building | State or Region | Disinfect. | Type | Hot Water Recirc. | Closure Date | Sampling Date(s) | Description of Occupancy/Closures | Preventative Measures During Low Occupancy Period | Measures after Study, Follow-Up, or Response Actions |
|---|---|---|---|---|---|---|---|---|---|---|
| MI | MI-1 | Michigan | Mono-chloramine | Full-Scale | Yes | 3/14/20 | 8/21/20 | Occupancy was restricted to essential personnel (~25%). | Closed water fountains. Random fixture flushing. Pool was being refilled weekly to flush water. | Newly developed flushing and building recommissioning guidelines. |
| | MI-2 | | | | Yes | 3/14/20 | 8/24/20 | | Closed water fountains. Cold water in the building was being flushed every two weeks. Building was last flushed on 8/4/20. | |
| | MI-3 | | | | Yes | 3/14/20 | 8/26/20 | | Building water was being sparsely used but not intentionally flushed or managed. | Resampled due to *L. pneumophila* detection. Reported water heater setpoint was 120F (49C), despite lower temps even after extended flushing. After detection, hot water tanks were drained. Full building flush conducted for 15 minutes on 9/5/20. Newly developed flushing and building recommissioning guidelines. |
| VA | VA-1 | Virginia | Mono-chloramine | Full-Scale | Yes | 3/16/20 | 7/26/20 | Occupancy was restricted to essential personnel. | One-time flushing event April (week of 4/20/2020). Opened most outlets for 1-3 minutes. | None |
| | VA-2 | | | | Yes | 3/16/20 | 7/28/20 | | | |
| MA | MA-1 | Massachusetts | Mono-chloramine | Full-Scale | Yes | 3/13/20 | 6/5/20 | Occupancy reduced to ~5% as of 3/23/20, phased reopening, starting June 2020 | Unsure. From the water usage data, the building water was sparsely used. Water usage increased by 4 logs with phased reopening, starting June | Maintenance activities (cleaning) likely in the building during the stagnation period, so there may have been minor water usage. |
| | MA-2 | | | | Yes | 3/13/20 | 6/5/20 | | | |
| | MA-3 | | | | Yes | 3/13/20 | 6/5/20 | | | |
| CA | CA-1 | California | Mono-chloramine | Full-Scale | Yes | 4/1/20 | 7/16/20 | Occupancy reduced to 2-4% in April, then 0% in June 2020 | No preventative measures were taken during the study period. | No preventative action within the buildings. Maintenance activities (WIFI repairs and cleaning) meant there may have been minor water usage |
| | CA-2 | | | | Yes | 4/1/20 | 7/16/20 | | | |
| | CA-3 | | | | Yes | 4/1/20 | 7/16/20 | | | |
| CH | CH-1 | Switzerland | None | Full-Scale | Yes | 3/9/20 | 4/24/20 | 5% occupancy from 3/9/20 to April 28, 2020; maximum 30% occupancy 4/28/20 through the end of the year | Swiss Federal Guidelines: Flush all fixtures until maximum temperature is reached. Did this one time before reopening the building. Emptied the boiler hot water during flushing several times. | None |

#### SI 1.2 – Sampling and Analysis

**Table S2.** Summary of total number of samples totals by site, flush condition, and type of analysis.

| | IN | OH | AZ | PA | WV | QC | MI | VA | MA | CA | CH | | n |
|---|---|---|---|---|---|---|---|---|---|---|---|---|---|
| All study samples | 12 | 4 | 7 | 18 | 30 | 56 | 19 | 18 | 12 | 20 | 62 | | 258 |
| First-draw | 11 | 4 | 7 | 9 | 19 | 56 | 19 | 9 | 6 | 20 | 43 | | 203 |
| Flushed | 1 | | | 9 | 11 | | | 9 | 6 | | 19 | | 55 |
| Legiolert | 12 | 4 | 7 | 18 | 30 | 56 | 19 | 18 | 12 | 20 | 62 | | 258 |
| qPCR | 4 | 4 | 7 | | 30 | 23 | 19 | 16 | | 17 | | | 120 |
| ddPCR | | | | 18 | | | | | | | | | 18 |
| Chlorine (total and/or free) | 2 | | 7 | 18 | 30 | 56 | 19 | 18 | 12 | 20 | | | 182 |
| Temperature | 2 | | 7 | 18 | 30 | 56 | 19 | 18 | 12 | 20 | | | 182 |
| pH | 2 | | 7 | 18 | 29 | 56 | 19 | 18 | 12 | 20 | | | 181 |
| | Free chlorine | | | | | | Chloramines | | | | None | | |

**Table S3.** Summary of sampling and analysis controls. A subset of these controls was analyzed by each site.

| Control | Description | Analysis |
|---|---|---|
| Trip Control | 1.1 L of autoclaved Milli-Q/Nanopure water that was kept in the cooler during sampling trips. | Legiolert and qPCR/ddPCR |
| Environmental Control | 1.3 L of autoclaved Milli-Q/Nanopure water that was brought to the site and opened to expose it to the site environment. | Legiolert and qPCR/ddPCR |
| Legiolert Reagents/Materials Negative Control | 100 mL of autoclaved Milli-Q/Nanopure water added to an unused sample container and processed with samples | Legiolert |
| Legiolert Kit Lot Negative Control | Per manufacturer instructions, *Enterococcus faecalis* ATCC 29212 performed at least once per kit lot | Legiolert |
| Legiolert Kit Positive Control | Per manufacturer instructions, *L. pneumophila* performed at least once per kit lot | Legiolert |
| Filtration Set-Up Control | Sterile water filtered using filtration set-up. | Legiolert and qPCR/ddPCR |
| Filter Control | Unused filter. | qPCR/ddPCR |
| DNA Extraction Control | Empty tube processed with samples. | qPCR/ddPCR |
| qPCR/ddPCR Negative Control | Reaction mix with sterile water instead of sample. | qPCR/ddPCR |
| qPCR/ddPCR Positive Control | Synthetic DNA (gBlock) | qPCR/ddPCR |

**Table S4.** Summary of physicochemical methods.

| | Free Chlorine | | Total Chlorine | | Monochloramine | | | | | |
|---|---|---|---|---|---|---|---|---|---|---|
| Site | Method | DL (mg/L as $Cl_2$) | Method | DL (mg/L as $Cl_2$) | Method | DL (mg/L as $Cl_2$) | pH | Temperature | Conductivity | Dissolved Oxygen |
| IN & OH | DPD method HACH Pocket Colorimeter DR300 | 0.02 | DPD method HACH Pocket Colorimeter DR300 | 0.02 | NA | | Oakton 450 pH probe | Oakton 450 pH probe | NA | YSI ProODO Optical Dissolved Oxygen Instrument |
| AZ | DR 900 colorimeter - program 80 - DPD 8021 method | 0.02 | DR 900 colorimeter - program 80 - DPD 8167 method | 0.02 | NA | | Oakton pH30 probe (pH30 pH tester) | Ryobi IR002 Infrared Thermometer | Thermo scientific - Orion Versa Star Pro - pH/ISE/Conductivity/ Dissolved Oxygen Multiparameter Benchtop Meter | |
| PA | DPD method- Hach Method 8021 | 0.02 | DPD Method - Hach Method 10250 | 0.05 | NA | | Hanna Combo Multiprobe | | NA | NA |
| WV | DPD method- Hach Method 8021 | 0.02 | NA | | NA | | Thermo Scientific Orion Star A326 Portable Meter | | | NA |
| QC | DPD method 8021, HACH DR 2800 portable spectrophotometer | 0.02 | DPD method 8167, HACH DR 2800 portable spectrophotometer | 0.02 | NA | | HACH HQ40d digital portable multi-probes meter | Digital thermometer | HACH HQ40d digital portable multi-probes meter | HACH HQ40d digital portable multi-probes meter |
| MI | DPD method- Hach Method 10245 | 0.05 | DPD Method - Hach Method 10250 | 0.05 | Indophenol Method- Hach Method 10200 | 0.04 | Hanna Instruments HI98121 portable probe | | NA | NA |
| VA | DPD method- Hach Method 8021 | 0.02 | DPD Method - Hach Method 10250 | 0.05 | NA | | Thermo Scientific Orion 110 Series meter with ATC | | NA | NA |
| MA | NA | | DPD Method - Hach Method 8167 | 0.02 | NA | | Thermo Scientific Orion Star A325 Multiparameter Meter | | | |
| CA | DPD method- Hach Method 8021 | 0.02 | DPD Method - Hach Method 8167 | 0.02 | NA | | Thermo Scientific Orion STAR A326 Portable Meter | | | |
| CH | NA | | | | | | | | | |

DL: Detection limit
NA: Analysis not performed.

**Table S5.** DNA collection, extraction, and quantification.

| Site | Filter Type | Sample Preservation and Storage | DNA Extraction Method | Sampling and Extraction Controls | DNA Quantification |
|---|---|---|---|---|---|
| IN & OH | 0.4 µm polycarbonate membrane filter disks filters (EMD Millipore, HTTP04700). | Filters aseptically transferred to microcentrifuge tubes and stored at -80˚C until DNA extraction. | DNeasy Power Water Kit (QIAGEN, 14900-100-NF) | An extraction negative control and a filter negative control were included for each extraction session. | NanoDrop |
| AZ | 0.2 µm polycarbonate membrane filter disks (EMD Millipore, GTTP04700) | Filters aseptically transferred to 2 mL microcentrifuge tubes and stored at -80˚C until DNA extraction. | DNeasy Power Soil Pro kit (QIAGEN, 47014). For bead beating - Precellys evolution which was set to 10,000 rpm, 3 cycles for 15 sec with 10 sec pause. | Trip, environmental (field blank), and filter negative controls collected during each sampling event. An extraction negative control was included for each extraction session. | Thermo Scientific NanoDrop 2000 spectrophotometer |
| PA | 0.2 µm polycarbonate membrane filter disks (EMD Millipore, GTTP04700) | Filters aseptically transferred to 2 mL sterile microcentrifuge tubes and stored at -20˚C until extraction | FastDNA SPIN kit (MP Biomedicals, 116540600-CF) with bead beating instead of the FastPrep step. | Trip, environmental, filter, and filtration set-up negative controls collected during each sampling event. An extraction negative control was included for each extraction session. | No DNA quantification |
| WV | 0.2 µm polycarbonate membrane filter disks (EMD Millipore, GTTP04700) | Filters aseptically transferred to 2 mL sterile screw top tubes and stored at -20˚C until extraction | FastDNA SPIN kit (MP Biomedicals, 116540600-CF) with bead beating instead of the FastPrep step. | Environmental and field negative controls included each sampling day. Filter control included in each extraction session. | No DNA quantification |
| QC | 0.2 µm polyethersulfone membrane filter disks (PALL Corporation, 66234) | Filters aseptically transferred to sterile microcentrifuge tubes and stored at -80˚C until DNA extraction (approx. 1-10 months) | 1) FastPrep Lysing Matrix A (MP Biomedicals, 116910050-CF) with FastPrep-24 bead beater (6 m/s, 40s, 2x) and centrifugation (13200 rpm, 5min, 1x), repeated overall 2x, 2) ammonium acetate impurities precipitation and centrifugation (13200 rpm, 15min, 4˚C, 2x), 3) overnight (4˚C) isopropanol DNA precipitation, 4) centrifugation (13200 rpm, 30min, 4˚C) and successive ethanol washes, 5) 100 µL sterile PCR buffer addition | No sampling or analysis controls. | No DNA quantification |

| Site | Filter Type | Sample Preservation and Storage | DNA Extraction Method | Sampling and Extraction Controls | DNA Quantification |
|---|---|---|---|---|---|
| MI | 0.2 µm polycarbonate membrane filter disks (EMD Millipore, GTTP04700) | Filters aseptically transferred to 2 mL sterile screw top tubes and stored at -80˚C until extraction | FastDNA SPIN kit (MP Biomedicals, 116540600-CF) with 2 minutes of bead beating with a Bio Spec Mini bead beater instead of the FastPrep step. | Trip, environmental, filter, and filtration set-up negative controls collected during each sampling event. An extraction negative control was included for each extraction session. Positive Legiolert controls per lot. A negative Legiolert control was included for each sampling event. | Qubit dsDNA High Sensitivity assay kit with a Qubit 2.0 fluorometer (Thermo Scientific) |
| VA | 0.2 µm polycarbonate membrane filter disks (EMD Millipore, GTTP04700) | Filters aseptically transferred to 2 mL sterile screw top tubes and stored at -20˚C until extraction | Filters frozen at VT; FastDNA Spin Kit with FastPrep Homogenization | Sampling negative control included during each sampling day. DNA extraction negative control (unused filter) included in each extraction session. Legiolert positive control (manufacturer supplied) and negative control (autoclaved tap water) per lot. | No DNA quantification |
| MA | NA | | | | |
| CA | 0.22 µm polyethersulfone cartridge filters (EMD Millipore, Z359912) | Filters aseptically transferred to 50 mL sterile screw top tubes and stored at -80˚C until extraction | Modified DNeasy Power Water Kit (QIAGEN, 14900-100-NF). Protocol detailed in dx.doi.org/10.17504/protocols.io.66khhcw. | Environmental and field controls included each sampling day.<br>Filter control included in each extraction batch | Qubit dsDNA High Sensitivity assay kit with a Qubit 4 fluorometer (Thermo Scientific) |
| CH | NA | | | | |

NA: Sample DNA not collected.

**Table S6.** qPCR and ddPCR primers, probes, and standards.

| Lab | Sites | Analysis Method | Assay | Gene Target | Amplicon Length (bp) | Forward Primer (5’-3’) | Reverse Primer (5’-3’) | Probe (5’-3’) | Standard Sequence (5’-3’) |
|---|---|---|---|---|---|---|---|---|---|
| A | VA, WV, IN, OH, CA | qPCR | Nazarian et al. 2008 | *mip* | 79 | LmipF: 5’- AAAGGCATGC AAGACGCTATG -3’ (21 bp, IDT) | LmipR: 5’- GAAACTTGTT AAGAACGTCT TTCATTTG-3’ (28 bp, IDT) | LmipP: 5’- FAM- TGGCGCTCA ATTGGCTTTA ACCGA-BHQ2-3’ (24 bp, IDT) | 5'-AGCTGTCAGCACTACTAACTT GCGGTCAGTAAAGGCATG CAA GAC GCT ATG AGT GGC GCT CAA TTG GCT TTA ACC GAA CAG CAA ATG AAA GAC GTT CTT AAC AAG TTT CTG CAT GAT CTA CGT GCG TCA CAT GCA GTA C-3' (139 bp, gBlock, IDT) |
| B | AZ | | | | | | | | |
| C | PA | ddPCR | Wullings et al., 2011 | *mip* | 120 | LpneuF: 5’-CC GATGCCACATC ATTAGC-3’ (19 bp, IDT) | LpneuR: 5’- CCAATTGAGC GCCACTC ATAG-3’ (21bp, IDT) | None | 5'-CCGATGCCACATCATTAG CTACAGACAAGG ATAAGTTGTCTTATAG CATTGGTGCCGATTTGGGGAAGAAT TTT AAAAATCAAGG CATAGATGTTAATCCGGAAGCAAT GGC TAAAGGCATGCAAGACGCTATGAG TGGCGCTCAATTGG-3' (150 bp, gBlock, IDT) |
| D | QC | qPCR | Bio-Rad's iQ-Check Quanti Lp real-time PCR kit (cat. no. 3578103) proprietary assay | | | | | | Proprietary kit standards |
| E | MI | qPCR | Nazarian et al. 2008 | *mip* | 79 | LmipF: 5’- AAAGGCATGC AAGACGCTAT G-3’ (21 bp, IDT) | LmipR: 5’- GAAACTTGT TAAGAACGT CTTTCATTTG -3’ (28 bp, IDT) | None | 5'-AGCTGTCAGCACTACTAACTT GCGGTCAGTAAAGGCA TG CAA GAC GCT ATG AGT GGC GCT CAA TTG GCT TTA ACC GAA CAG CAA ATG AAA GAC GTT CTT AAC AAG TTT CTG CAT GAT CTA CGT GCG TCA CAT GCA GTA C-3' (139 bp, gBlock, IDT) |

**Table S7.** qPCR and ddPCR protocols.

| Lab | Method | Master Mix | Instrument | Std curve range | Reaction Mix | Cycling conditions | Replicates |
|---|---|---|---|---|---|---|---|
| A | qPCR with Nazarian et al. 2008 | 2X SsoFast Probes Supermix (Bio-Rad, cat no. 1725230) | Bio-Rad CFX96 real-time | 5 – $10^7$ gc/rxn | 10 µL reactions: 5 µl of master mix, 250 nM of forward and reverse primers, 93.75 nM probe, and 1 µL of DNA template. | 95°C for 2 min, 40 cycles of 95°C for 5 s and 60°C for 10 s | 3x |
| B | qPCR with Nazarian et al. 2008 | SSO Fast EvaGreen (Bio-Rad, cat no. 1725200) | Bio-Rad CFX96 real-time | 30 – $10^7$ gc/rxn | 25 µL total reaction volume: 12.5 µL universal probe mix, 1.25 µL - 10 µM forward primer (final conc 500 nM), 1.25 µL - 10 µM reverse primer (final conc 500 nM), 0.6 µL - 10 µM probe (final conc 250 nM), 6.4 µL water, 3 µL DNA template | 95°C for 2 min, 40 cycles of 95°C for 5s, 60°C for 30s, 72°C for 30s | 3x |
| C | ddPCR with Wullings et al., 2011 | EvaGreen Supermix (Bio-Rad, cat no. 1864034) | Bio-Rad ddPCR QX200 Droplet Generator, C1000 Touch thermocycler, QX200 Droplet Reader | N/A | 22 µL reactions: 11 µL of master mix, 0.44 µL of 10 µM forward and reverse primers (final conc 0.2 µM), 0.55 µL of 50 mg/mL BSA (Invitrogen, final conc 0.625 mg/mL, 2 µL DNA template, 7.57 µL water | 95°C for 5 min, 45 cycles of 95°C for 30 s, 57°C for 1 min, 72°C for 1 min, 4°C for 5 min, 90°C for 5 min | N/A |
| D | qPCR Bio-Rad's iQ-Check Quanti Lp real-time PCR kit (cat. no. 3578103) | | Rotor-Gene Q QIAGEN | $10^1$ – $10^4$ gc/rxn | 50 µL total rxn volume: 45 µL amplification mix, 5 µL extracted DNA in sterile PCR buffer | 95°C for 15 min, 50 cycles of 95°C for 15s, 57°C for 30s, 72°C for 30s, 72°C for 15 min | 2x |
| E | qPCR with Nazarian et al. 2008 | Fast EvaGreen w/ low ROX (2x, Biotium, cat. no. 31014) | Applied Biosciences QuantStudio 3 | $10^1$ – $10^8$ gc/rxn | 10 µL total rxn volume: 5 µL of master mix, 0.5 µL of 10 µM forward and reverse primers (final concentration 0.2 µM), 0.625 µL of 25 mg/mL BSA (Invitrogen, final concentration 0.625 mg/mL), 3.25 µL water, and 1 µL DNA template | 95°C for 2 min, 40 cycles of 95°C for 5s, 60°C for 30s, 72°C for 30s | 3x |

#### SI 2 – qPCR and ddPCR Quality Assurance and Quality Control

**Table S8.** qPCR/ddPCR standard curve parameters.

| Site | Plate | Y-Intercept | Efficiency (%) | R Squared |
|---|---|---|---|---|
| PA | 1 | ddPCR | | |
| | 2 | | | |
| | 3 | | | |
| | 4 | | | |
| MI | 2 | 36.3 | 97.6 | 0.994 |
| | 3 | 38.0 | 89.6 | 0.999 |
| | 6 | 36.8 | 97.4 | 0.999 |
| | 7 | 37.2 | 97.9 | 0.999 |
| | 9 | 37.7 | 90.8 | 0.997 |
| | 10 | 39.1 | 87.4 | 0.994 |
| AZ | 1 | 39.9 | 98.9 | 0.998 |
| QC | 1 | 36.6 | 115.0 | 0.992 |
| | 2 | 37.4 | 115.0 | 0.995 |
| CA & WV (run by Lab A) | 6 | 44.3 | 83.8 | 0.982 |
| | 8 | 44.4 | 83.6 | 0.980 |
| IN & OH (run by Lab A) | 7 | 43.9 | 85.8 | 0.988 |
| Nonquantitative plates | | | | |
| VA | 1 | 50.3 | 62.6 | 0.970 |
| CA & WV (run by Lab A) | 1 | 44.7 | 75.1 | 0.982 |
| | 2 | 46.3 | 69.2 | 0.986 |
| | 3 | 43.9 | 93.4 | 0.935 |
| | 4 | 44.6 | 86.6 | 0.940 |

**Table S9.** qPCR/ddPCR limit of detection (LOD) and lower limit of quantification (LLOQ) testing results in gene copies per reaction (gc/rxn).

| Site | Run By | LOD (gc/rxn) | LLOQ (gc/rxn) |
|---|---|---|---|
| AZ | Lab B | 30 | 30 |
| QC (plate 1) | Lab D | 1.0 | 16.9 |
| QC (plate 2) | | 1.0 | 19.7 |
| MI | Lab E | 10.2 | 20.4 |
| VA | Lab A | 100 | 100 |
| CA | | | |
| WV | | | |
| IN & OH | | | |
| PA | Lab C | 6.1 | 6.1 |

##### Conversion of LOD and LLOQ to gene copies per liter

qPCR/ddPCR LOD and LLOQ were converted to gene copies per liter on a per sample basis, as shown in Equation S1.

**Equation S1.** Conversion of gene copies per reaction to per liter.

$$\frac{gc}{rxn} \times \frac{rxn}{template\ vol\ (\mu L)} \times \frac{dilution\ factor}{1} \times \frac{extraction\ elution\ vol\ (\mu L)}{sample\ vol\ (mL)} \times \frac{1{,}000\ mL}{L} = \frac{gc}{L}$$

#### <u>SI 3 – qPCR and ddPCR Inhibition Testing Summary</u>

All laboratories performing qPCR analyses were instructed to perform inhibition testing by analyzing a subset of samples at multiple dilutions. Execution of inhibition testing varied by laboratory and is summarized below.

**Laboratory A**

Laboratory A analyzed samples from Sites IN, OH, WV, VA, and CA. A subset of samples (n=12) were run undiluted as well as diluted at 1:4, 1:10, and 1:20 to assess inhibition. All samples were then processed at a 1:10 dilution based on the dilution that resulted in the highest concentrations during inhibition testing.

**Laboratory B**

Laboratory B analyzed samples from Site AZ. All samples were tested for the 16S rRNA target to confirm the extraction process was successful and to determine if samples were inhibited. Both undiluted and 1:10 diluted samples were tested for the 16S rRNA gene target, and the difference in their Cq was calculated. If the Cq difference did not fall in the range of 2 to 4 Cq, the samples were considered to have PCR inhibition. Samples with PCR inhibitors were subjected to a 10-fold dilution while testing for the *Legionella mip* gene target. All other samples were tested without any dilution with a full standard curve for each plate.

**Laboratory C**

Laboratory C analyzed samples from Site PA using ddPCR. Although ddPCR is less susceptible to inhibitors than qPCR, reactions included 0.625 mg/mL BSA to minimize inhibition. No separate inhibition testing was conducted, and samples were processed undiluted.

**Laboratory D**

Laboratory D analyzed samples from Site QC. For QC-3 building, all samples (including controls and standards) from plate 1 (n=16) and plate 2 (n=7) were tested for qPCR inhibition according to Bio-Rad's iQ-Check Quanti *L. pneumophila* real-time PCR kit (cat. no. 3578103) user guide. Briefly, inhibition was considered if the Cq sample is higher than the addition of the standards (n=4) mean and three times their standard deviation (Cq > mean Cq QS + 3*sigma). Among all samples, no inhibition was detected during the amplification process.

**Laboratory E**

Laboratory E analyzed samples from Site MI. Of the 19 samples (excluding controls) analyzed using qPCR, all were analyzed undiluted and at least one dilution. If inhibition was observed based on the delta Cq between the undiluted samples and the first dilution, additional dilutions were performed. Dilutions used included 1:2, 1:5, 1:10, 1:100, 1:1,000, and 1:10,000. The majority of samples did not amplify at any dilution.

#### SI 4 – qPCR and ddPCR Cross-Laboratory Validation Summary

Cross-laboratory validation was conducted at the laboratories using the Nazarian et al. assay (Laboratories A, B, and E). Cross-laboratory validation was performed using a synthetic DNA standard, which consisted of the Nazarian et al. amplicon (79 base pair [bp] with 30 bp neutral adaptors on both ends) at $10^9$ copies per microliter (µL, Integrated DNA Technologies [IDT], Coralville, IA, USA). The gBlock was ordered from IDT pre-eluted in nuclease-free water. Upon receipt, the gBlock was divided into 20 µL aliquots, which were frozen at -80°C, then shipped overnight on ice to participating labs. Each laboratory analyzed serial dilutions of the standard to $10^3$ gc/µL or $10^1$ gc/µL on the same plate as the laboratory's typical standard material. The universally quantified standards were all quantified within the tolerance range such that concentrations can be compared across laboratories.

**Figure S2.** Cross-laboratory validation results for the laboratories using the Nazarian et al. (2008) qPCR assay.

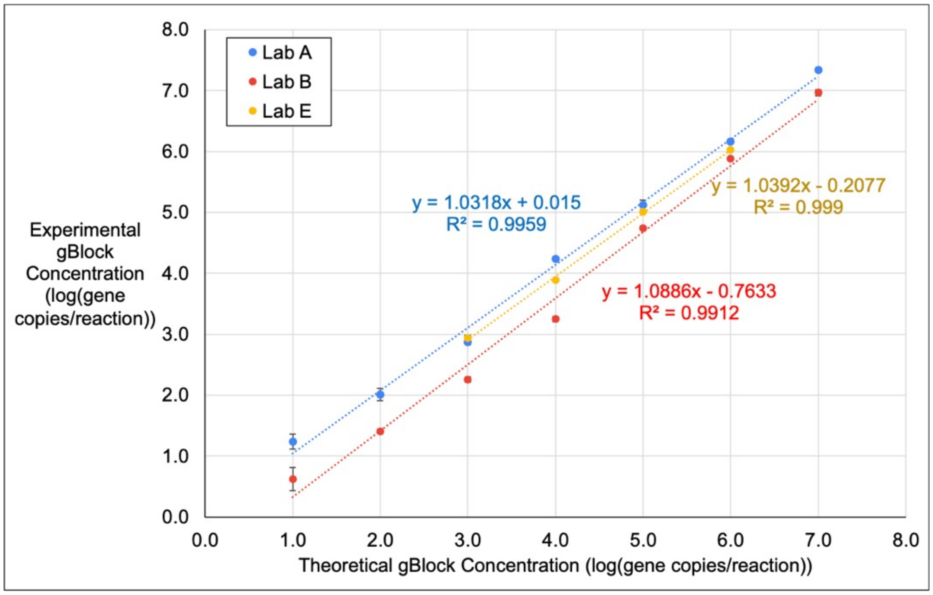

#### SI 5 – Results Supplement

**Table S10.** Physicochemical results summary.

| Parameter | Median Result (Count) | | | | | |
|---|---|---|---|---|---|---|
| | All Samples | By Disinfectant | | | By *L. pneumophila* Culture Result | |
| | | Free Chlorine | Mono-chloramine | None | *L. pneumophila* Positive | *L. pneumophila* Negative |
| Chlorine Residual (mg/L as $Cl_2$) | 0.05 (n=182) | <0.05 (n=113) | 0.28 (n=69) | --- | <0.05 (n=46) | 0.05 (n=136) |
| pH | 8.4 (n=181) | 8.2 (n=112) | 9.1 (n=69) | --- | 8.6 (n=46) | 8.3 (n=135) |
| Temperature (°C) | 25 (n=182) | 26 (n=113) | 23 (n=69) | --- | 29 (n=46) | 23 (n=136) |
| Dissolved Oxygen (mg/L) | 8.1 (n=90) | 7.0 (n=58) | 9.0 (n=32) | --- | 6.5 (n= 21) | 8.6 (n=69) |
| Electrical Conductivity (µS/cm) | 288 (n=95) | 300 (n=63) | 100 (n=32) | --- | 301 (n=27) | 276 (n=68) |

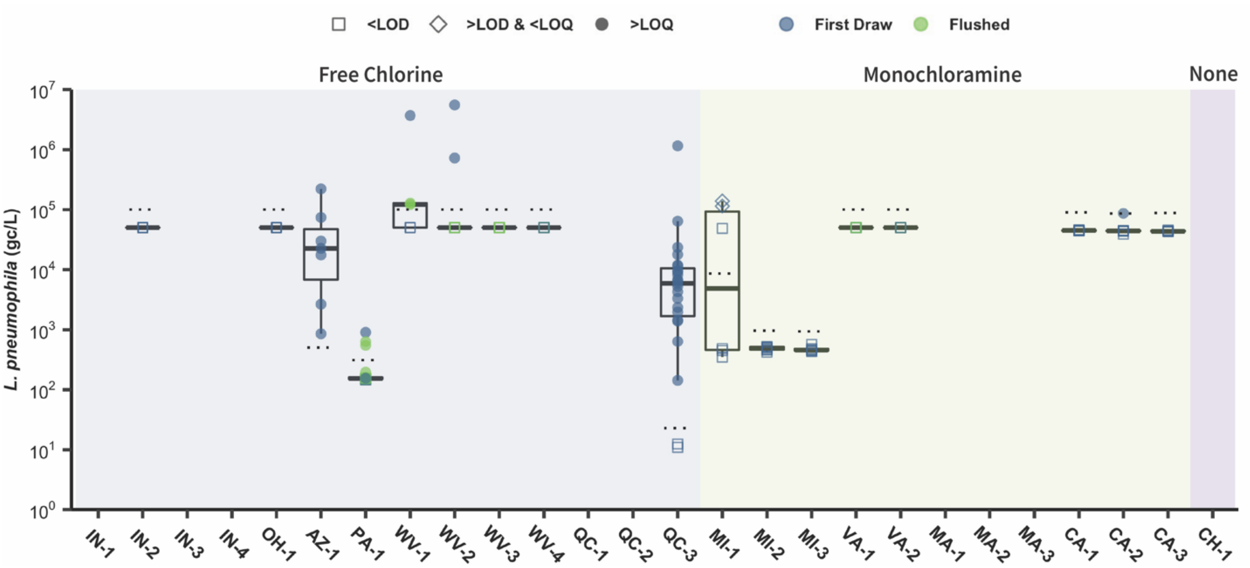


**Figure S3.** *L. pneumophila* qPCR/ddPCR results by building and secondary disinfectant type. Marker color represents sample type, where blue circles are first-draw samples and green circles are flushed samples. Results below the LOD are plotted at one-half the LOD and shown as open squares. Results above the LOD but below the LLOQ are shown as open diamonds. Results above the LLOQ are plotted as filled circles. LOD and LLOQ thresholds vary by laboratory depending on qPCR/ddPCR sensitivity (Table S9) and concentration/extraction volumes (Table S5). Dotted horizontal lines show the geometric mean of the LOD for each building.

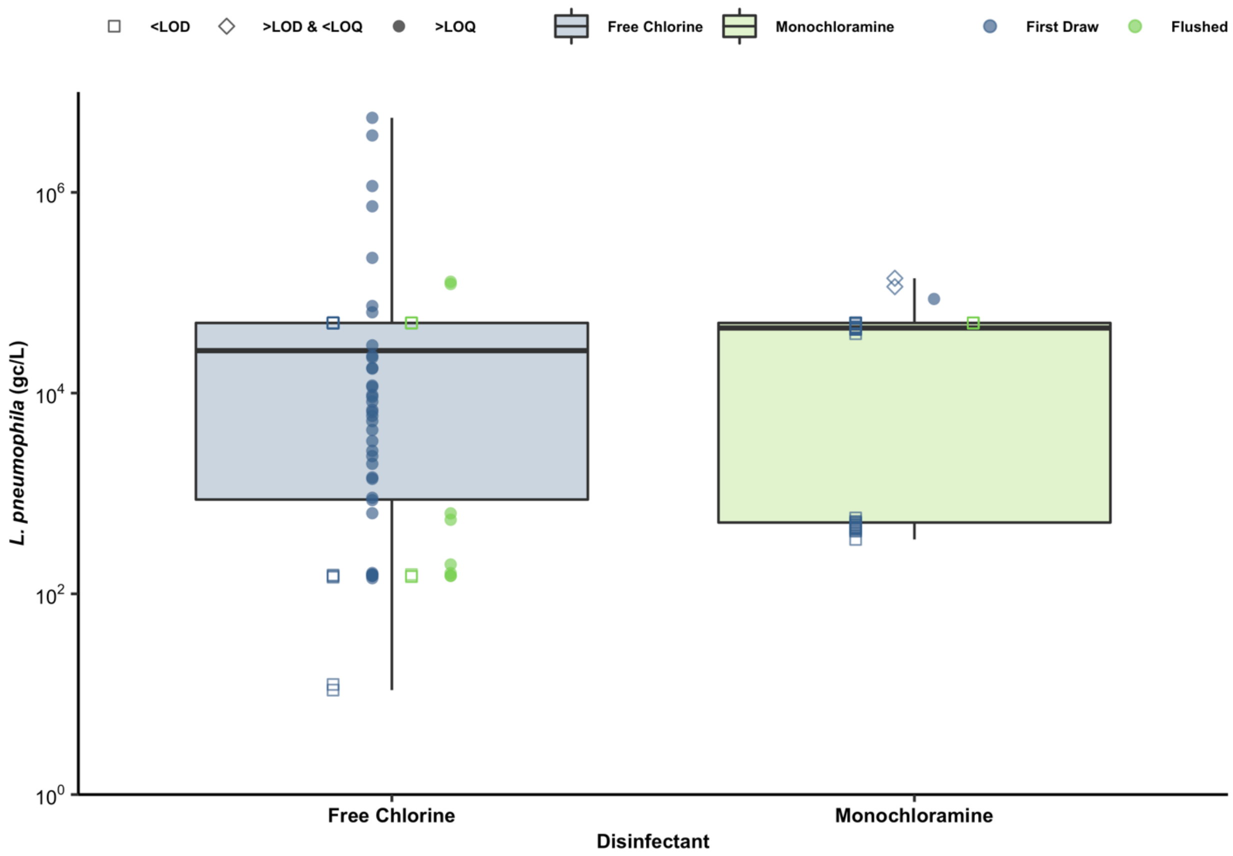


**Figure S4.** *L. pneumophila* qPCR/ddPCR results by disinfectant type and sample type. Marker color represents sample type, where blue circles are first-draw samples and green circles are flushed samples. Results below the LOD are plotted at one-half the LOD and shown as open squares. Results below the above LOD but below the LLOQ are shown as open diamonds. Results above the LLOQ are plotted as filled circles.

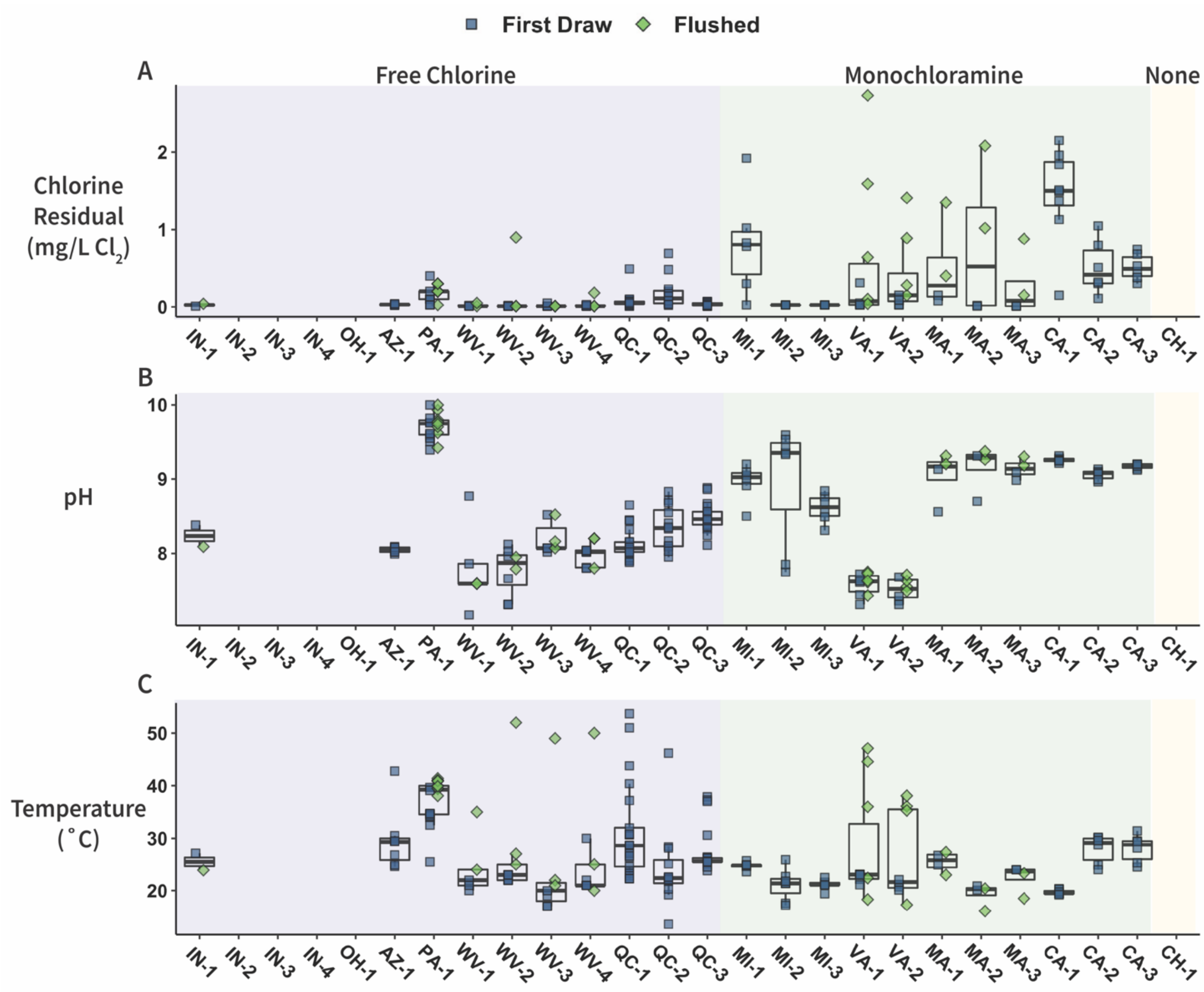


**Figure S5.** A) Chlorine residual, B) pH, and c) temperature by building and sample type. Marker color represents sample type, where blue squares are first-draw samples and green diamonds are flushed samples. Background colors represent disinfectant type: free chlorine, chloramine, or none. Temperature and chlorine were not measured for samples collected from Site CH.

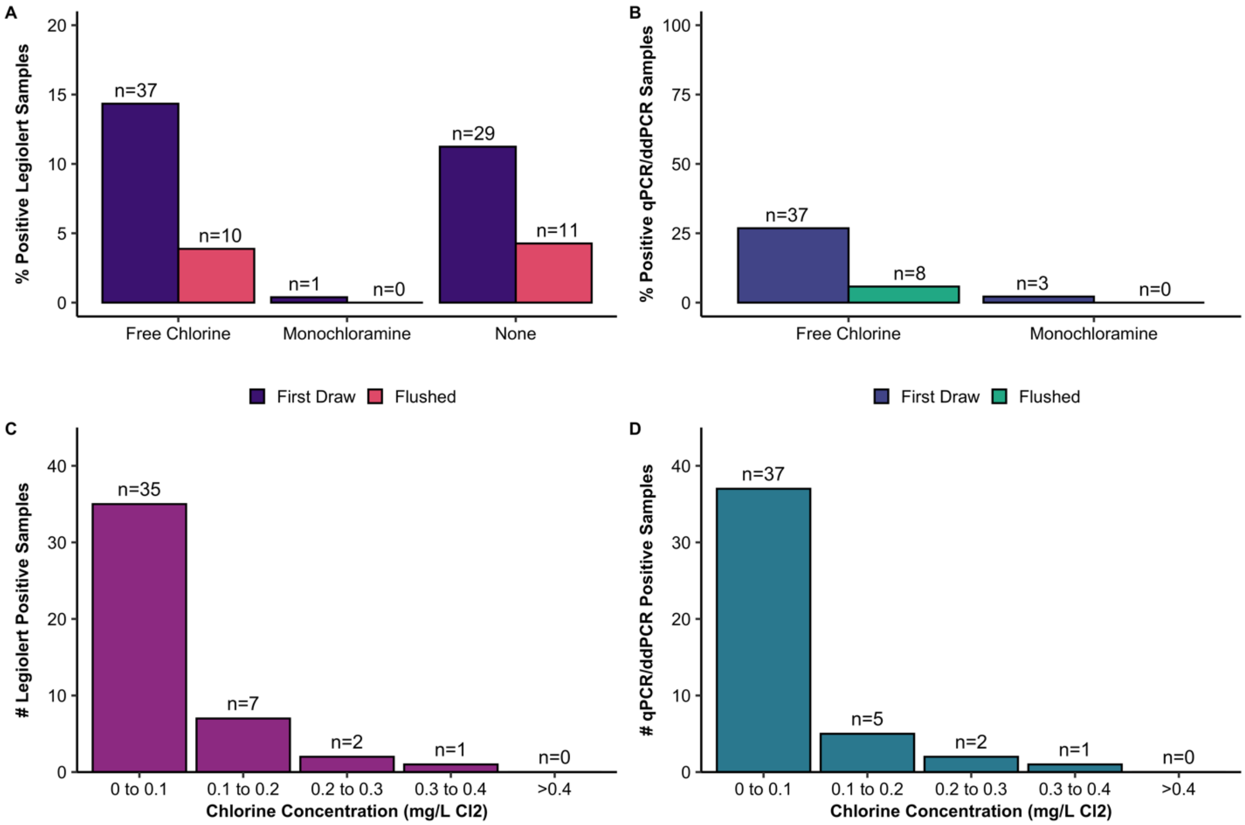


**Figure S6.** *L. pneumophila* Legiolert and qPCR/ddPCR results by disinfectant type and sample type. A) Percent Legiolert-positive samples by disinfectant type and flush condition, B) percent positive qPCR/ddPCR samples by disinfectant type and flush condition, C) number of Legiolert-positive samples by chlorine concentration **for free chlorine first draw and flushed samples only**, D) number of qPCR/ddPCR-positive samples by chlorine concentration **for free chlorine first draw and flushed samples only**.

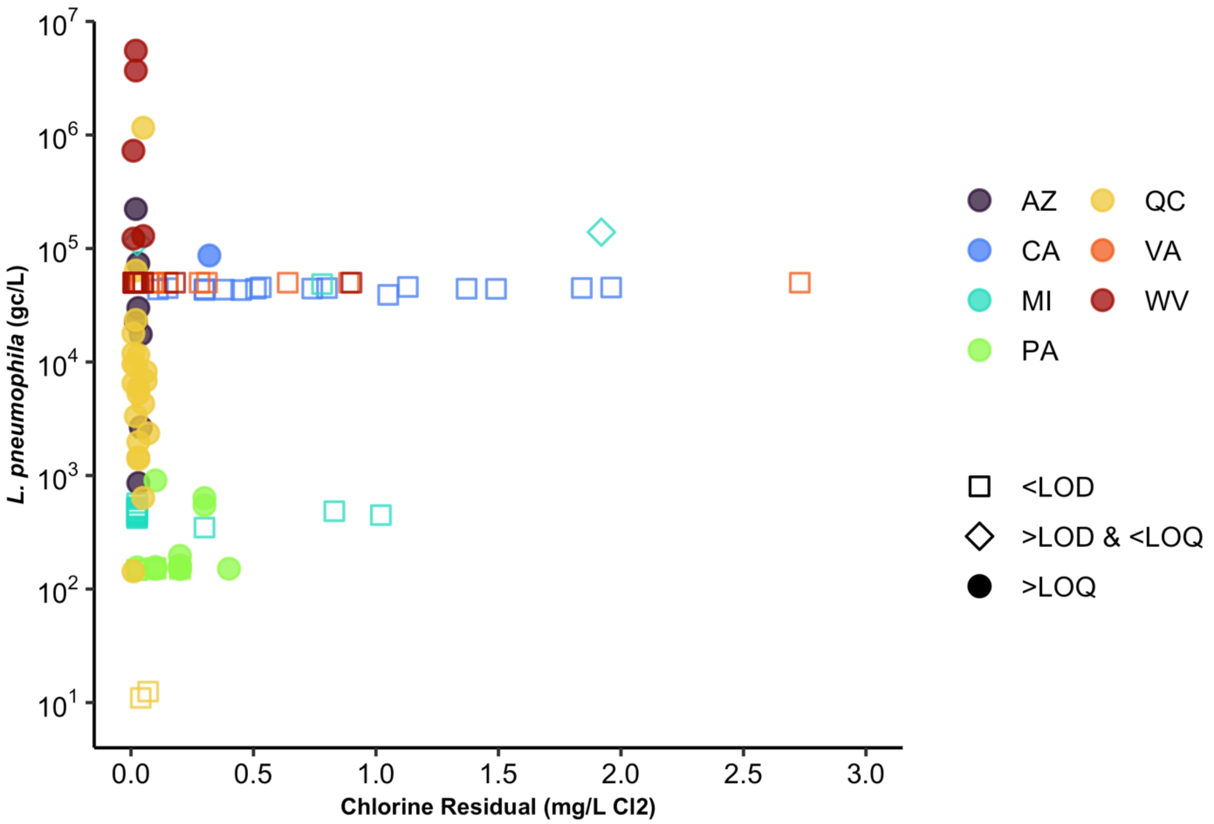


**Figure S7.** *L. pneumophila* qPCR/ddPCR results as a function of sample chlorine residual. Results below the LOD are plotted at one-half the LOD and shown as open squares. Results below the above LOD but below the LLOQ are shown as open diamonds. Results above the LLOQ are plotted as filled circles.

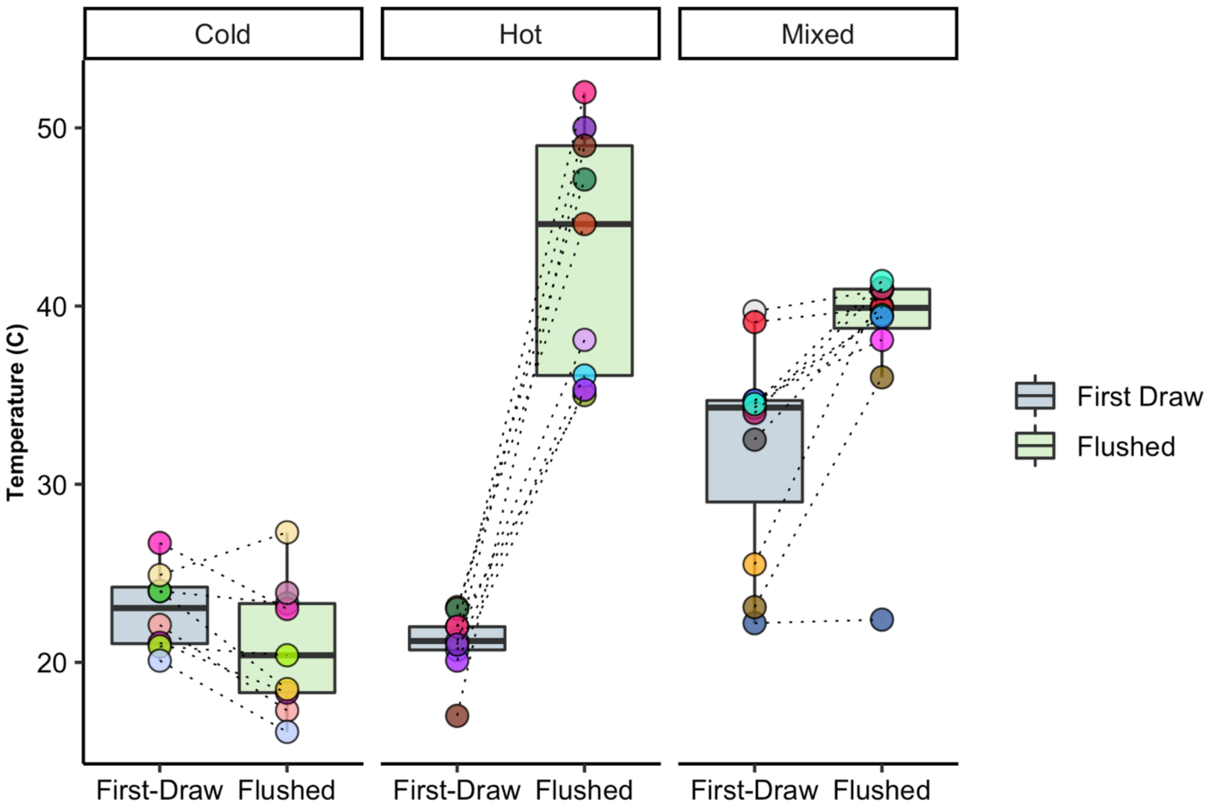


**Figure S8.** Sample temperature results by condition (first-draw vs. flushed) and fixture temperature (cold, hot, or mixed) **for paired samples.** Each point is a sample, and point color represents fixture identity.

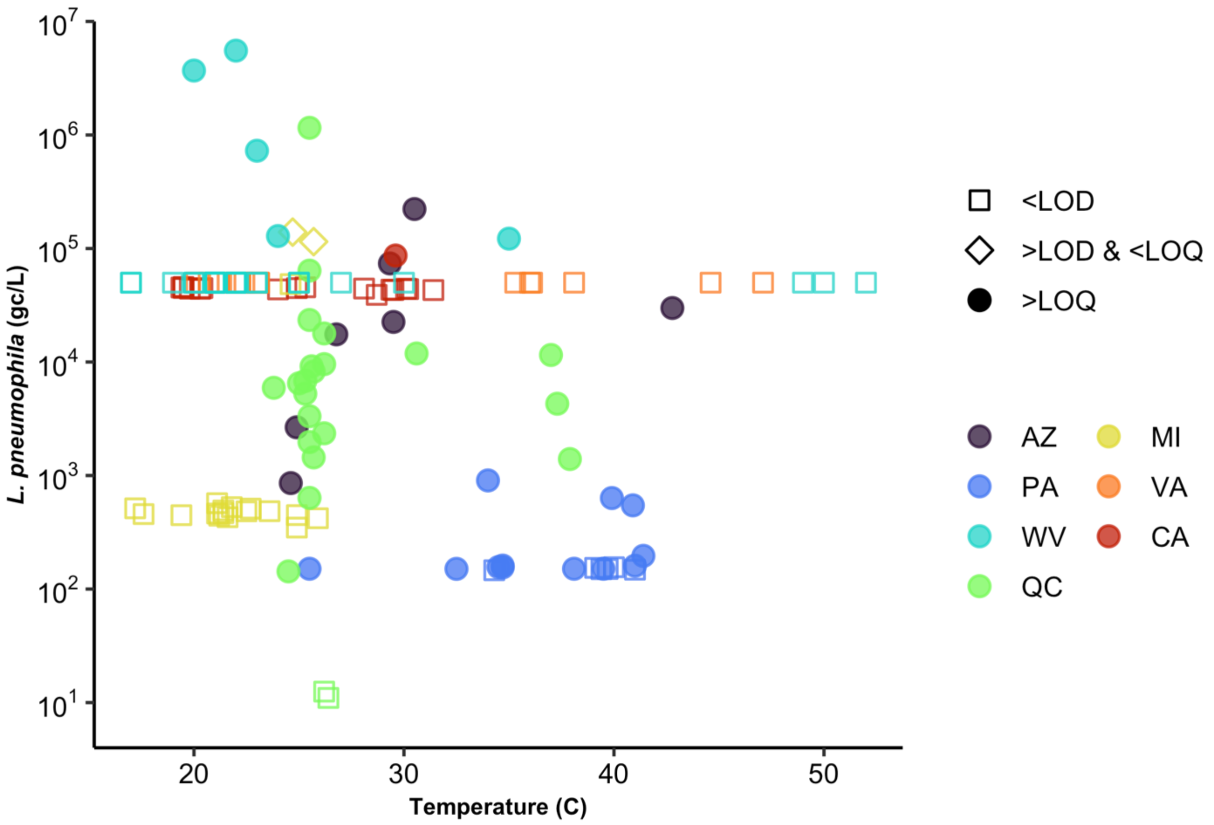


**Figure S9.** *L. pneumophila* qPCR/ddPCR results as a function of sample temperature. Results below the LOD are plotted at one-half the LOD and shown as open squares. Results above the LOD but below the LLOQ are shown as open diamonds. Results above the LLOQ are plotted as filled circles.

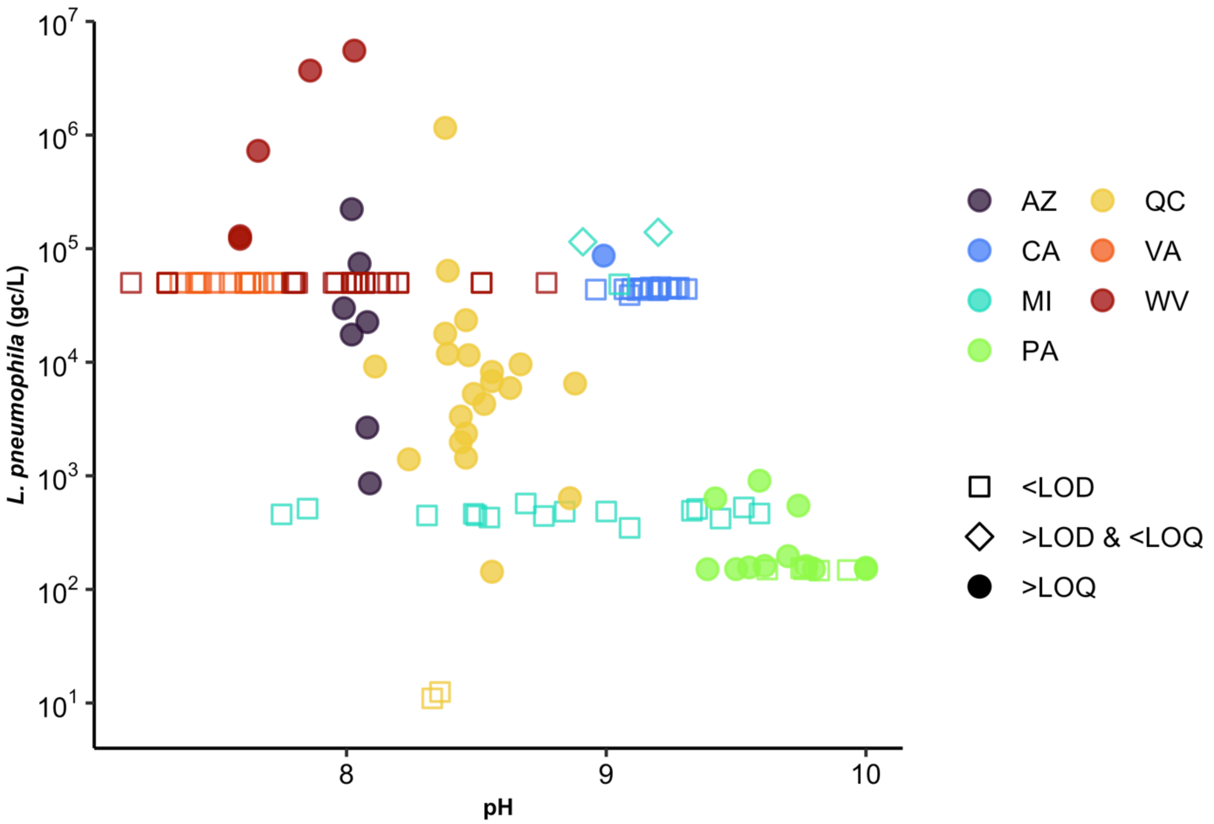


**Figure S10.** *L. pneumophila* qPCR/ddPCR results as a function of sample pH. Results below the LOD are plotted at one-half the LOD and shown as open squares. Results above LOD but below the LLOQ are shown as open diamonds. Results above the LLOQ are plotted as filled circles.

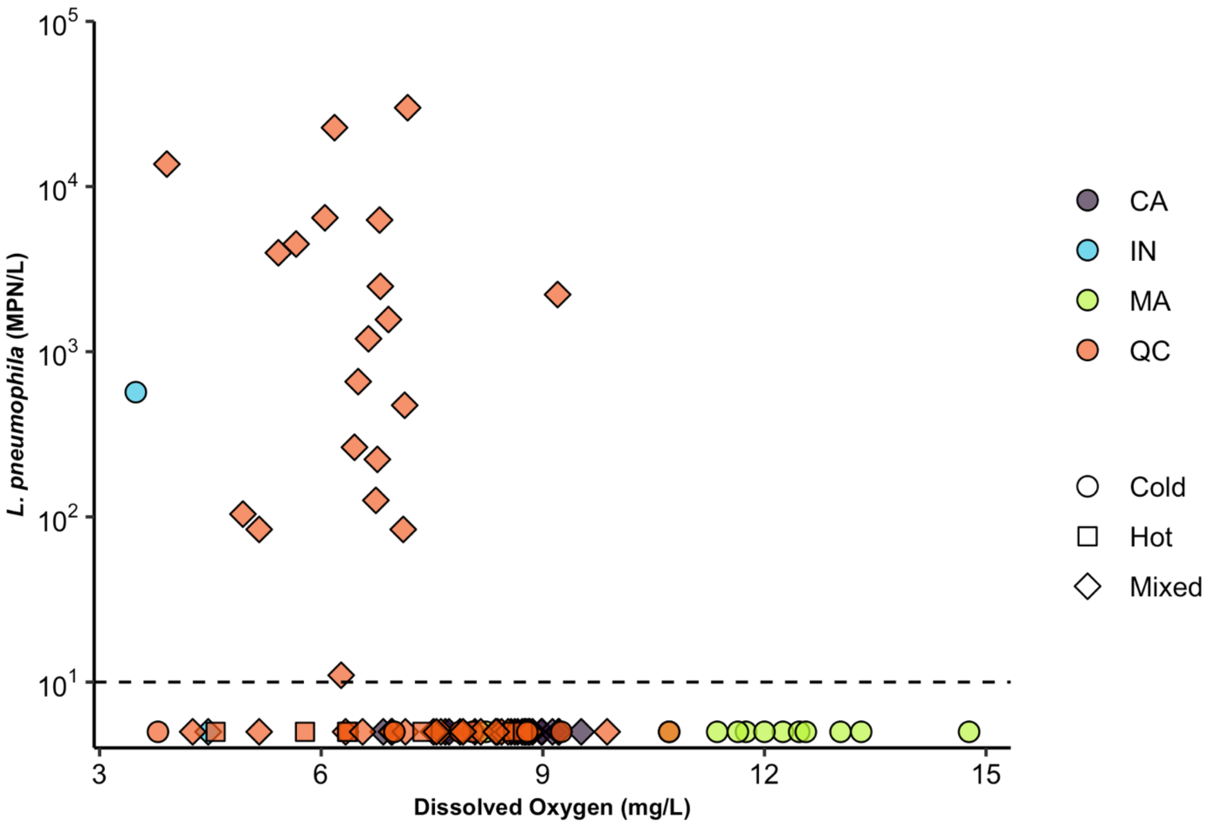


**Figure S11.** Dissolved oxygen as a function of culturable *L. pneumophila.*

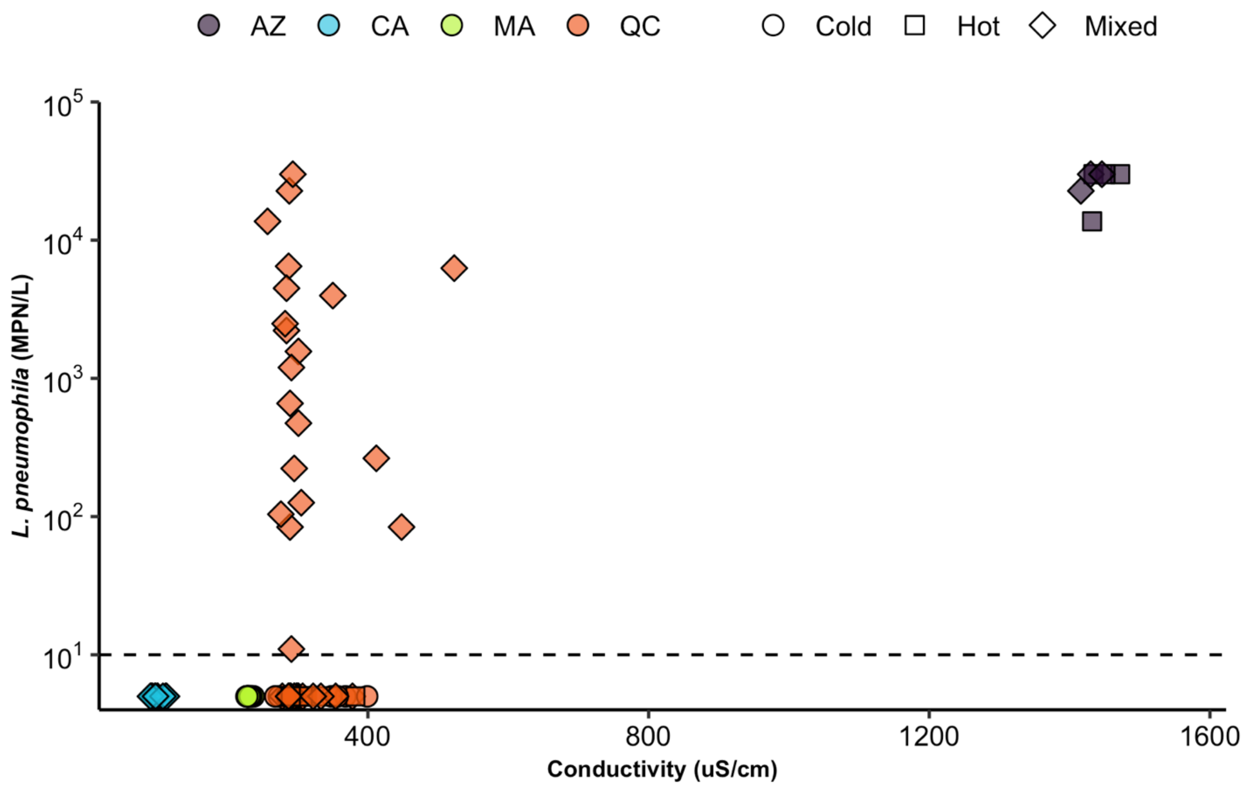


**Figure S12.** Electrical conductivity as a function of culturable *L. pneumophila.*

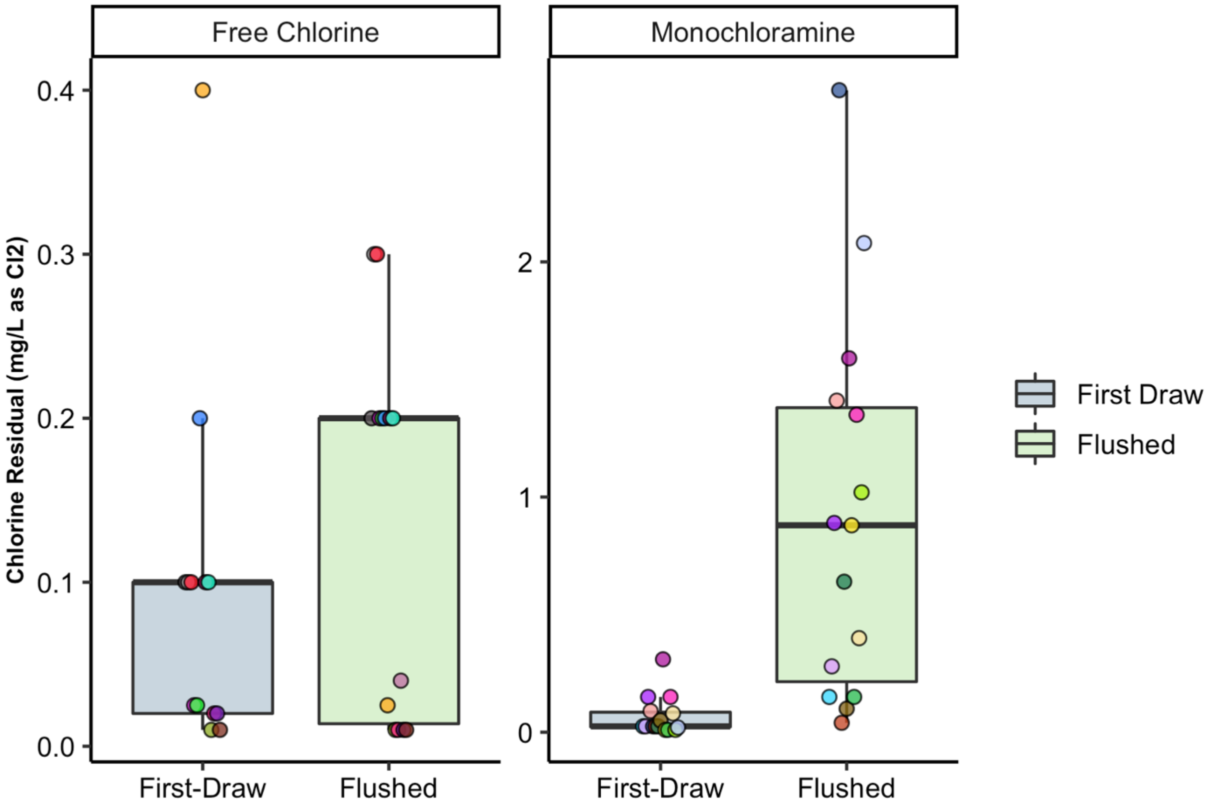


**Figure S13.** Sample chlorine residual results by condition (first-draw vs. flushed) and disinfectant type (free chlorine and chloramine) **for paired samples**. Points are colored by fixture.

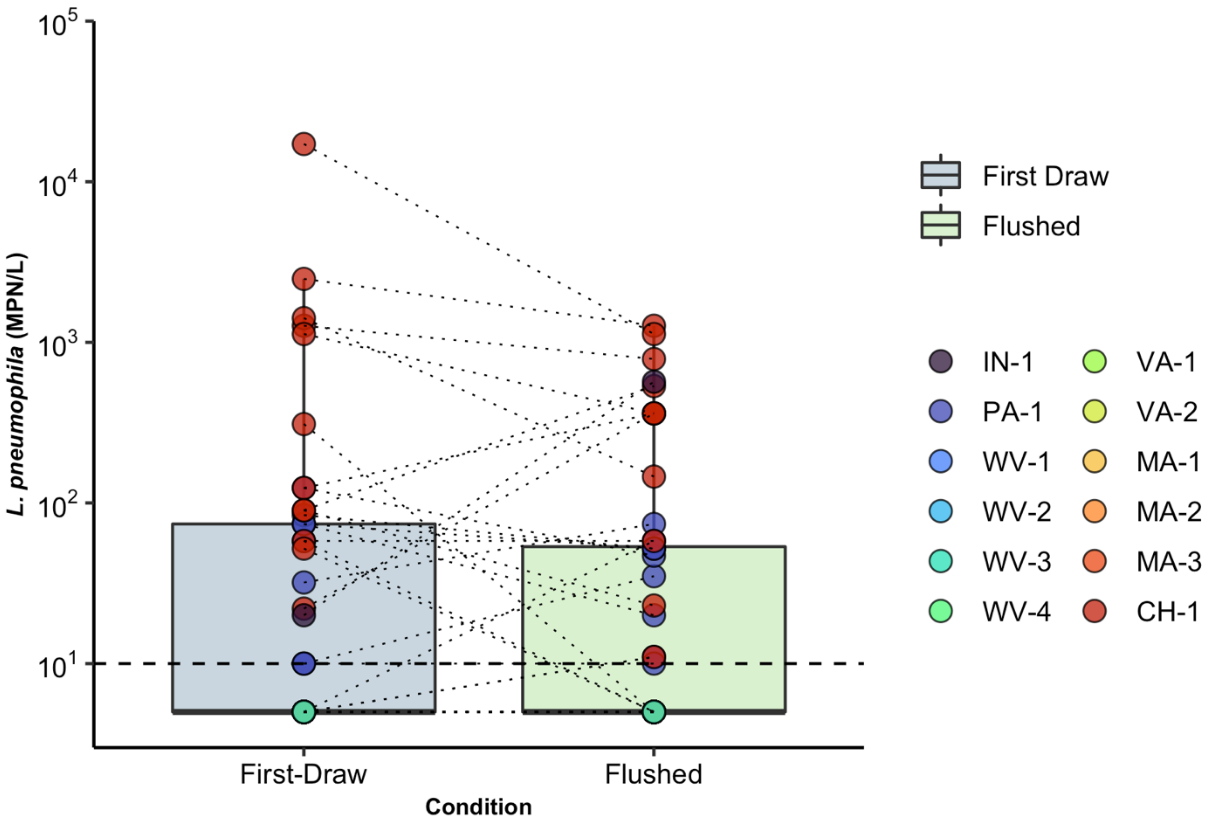


**Figure S14.** *L. pneumophila* Legiolert results (MPN/L) by condition (first-draw vs. flushed) **for paired samples**. Points are colored by building. The dashed line represents the LOD.

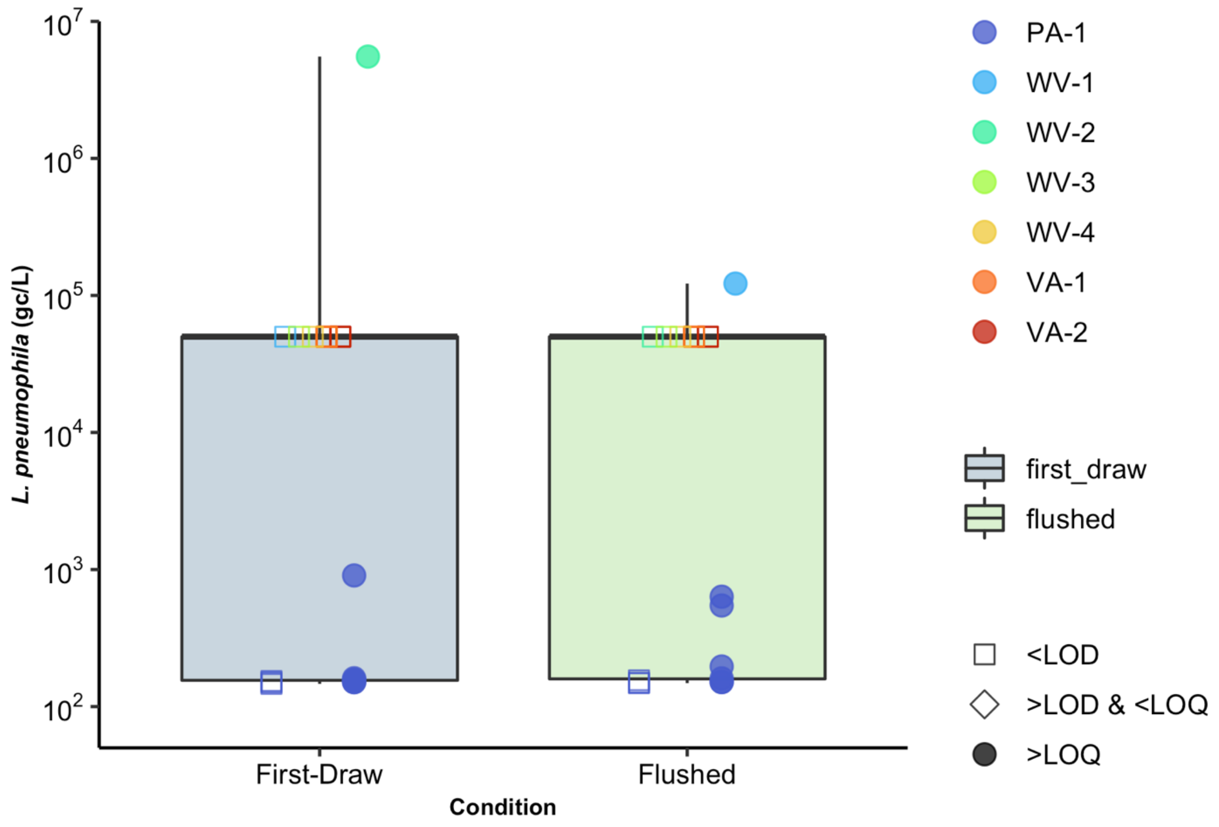


**Figure S15.** *L. pneumophila* qPCR/ddPCR results (gc/L) by site **for paired samples**. Results below the LOD are plotted at one-half the LOD and shown as open squares. Results above the LOD but below the LLOQ are shown as open diamonds. Results above the LOQ are plotted as filled circles. Points are colored by building.

**Input:**

```
output<-glmer(formula = lp_pos ~cl_tot_mgl + building_age+ temp_c+
pH+(1|building_id), data=glm1, family=binomial)
```

where

lp_pos: binomial vector where 0= *L. pneumophila* culture-negative and 1= *L. pneumophila* culture-positive

cl_tot_mgl: total chlorine result in mg/L as $Cl_2$

building_age: age of the building in years

temp_c: sample temperature in degrees Celsius

pH: sample pH

building_id: the unique identification name assigned to each building

**Output:**

```
Generalized linear mixed model fit by maximum likelihood (Laplace Approximation) [glmerMod]
 Family: binomial  ( logit )
Formula: lp_pos ~ cl_tot_mgl + building_age + temp_c + pH + (1 | building_id)
   Data: glm1

     AIC      BIC   logLik deviance df.resid
    74.4     90.7    -31.2     62.4      106

Scaled residuals:
    Min      1Q  Median      3Q     Max
-2.7575 -0.1778 -0.0705  0.2500  5.6366

Random effects:
 Groups      Name        Variance Std.Dev.
 building_id (Intercept) 11.48    3.388
Number of obs: 112, groups:  building_id, 10

Fixed effects:
              Estimate Std. Error z value Pr(>|z|)
(Intercept)  -21.66084   14.70340  -1.473    0.141
cl_tot_mgl    -0.29197    4.90055  -0.060    0.952
building_age  -0.03532    0.04002  -0.882    0.378
temp_c         0.03244    0.06311   0.514    0.607
pH             2.38928    1.63570   1.461    0.144

Correlation of Fixed Effects:
            (Intr) cl_tt_ bldng_ temp_c
cl_tot_mgl  -0.077
building_ag -0.366  0.082
temp_c      -0.182 -0.047  0.084
pH          -0.980  0.041  0.263  0.054
```

**Figure S16.** Generalized linear mixed effects model input and results for free chlorine samples with associated physicochemical measurements and building characteristics.

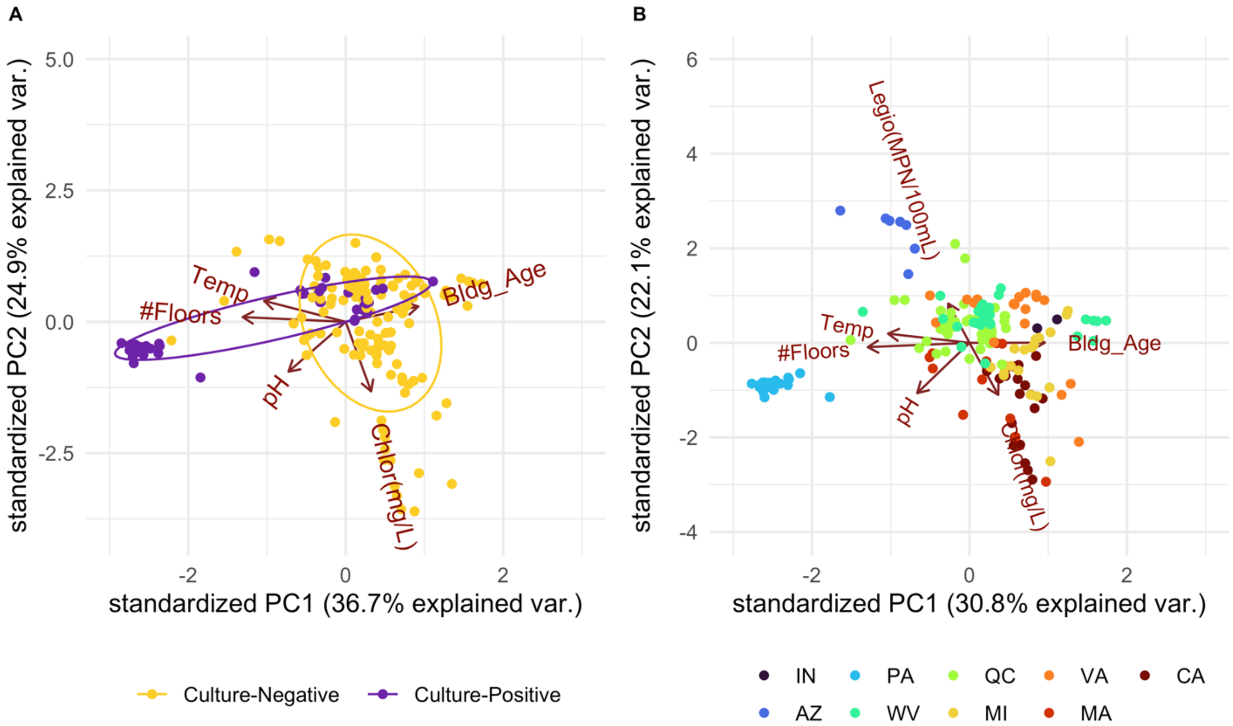


**Figure S17.** Principal component analysis (PCA) incorporating physicochemical parameters (chlorine residual, temperature, and pH) and building characteristics (number of floors and building age). A) Samples colored by culture-positivity or -negativity. B) Samples colored by site and axes include Legiolert concentration of *L. pneumophila*

Chlorine concentrations explained much of the variance in negative samples (A). Site PA clustered separately from the other sites, possibly because it was a separate replicated experimental system. Most of the variance in Site AZ was explained by *L. pneumophila* culture concentrations; whereas variance in other sites were mostly explained by other factors (B).
